# Validation of remote collection and quantification of blood Neurofilament light in neurological diseases

**DOI:** 10.1101/2023.12.04.23299336

**Authors:** Annabelle Coleman, Alexiane Touzé, Mena Farag, Marta Pengo, Michael J Murphy, Yara Hassan, Olivia Thackeray, Kate Fayer, Sophie Field, Mitsuko Nakajima, Elizabeth L Broom, Brook Huxford, Natalie Donkor, Ellen Camboe, Kamalesh C Dey, Alexandra Zirra, Aisha Ahmed, Ana Rita Gameiro Costa, Harriet Sorrell, Luca Zampedri, Vittoria Lombardi, Charles Wade, Sean Mangion, Batoul Fneich, Amanda Heslegrave, Henrik Zetterberg, Alastair Noyce, Andrea Malaspina, Jeremy Chataway, Sarah J Tabrizi, Lauren M Byrne

## Abstract

Promising blood-based biomarkers of neuropathology have emerged with potential for therapeutic development and disease monitoring. However, these tools will require specialist tertiary services for integration into clinical management. Remote sampling for biomarker assessment could ease the burden of in-person clinical visits for such tests and allow for frequent sampling. Here we evaluated a capillary finger-prick collection for remote quantification of blood neurofilament light (NfL), a common blood-based biomarker evident in various neurological disorders, and other exploratory markers of neuronal injury and neuroinflammation (GFAP, tau).

Matched samples from venepuncture and finger-prick were collected and processed into plasma and/or serum to directly compare NfL levels across four different neurological conditions (HD, MS, ALS, PD). Two delayed processing conditions were compared, three- and seven-day delay, simulating ambient shipment.

Capillary NfL and GFAP concentrations were equivalent to those in venous blood serum and plasma. Only NfL remained stable after seven-day processing delay. Capillary NfL replicated disease group differences displayed in venous blood.

This data supports our finger-prick method for remote collection and quantification of NfL. With the widespread applications for NfL across the spectrum of neurological disorders, this has the potential to transform disease monitoring, prognosis, and therapeutic development within clinical practice and research.

Graphical abstract: Figure 1

## Introduction

Blood-based biomarkers with pathobiological relevance to the central nervous system (CNS) have emerged over recent years with strong potential for clinical applications (1–4). However, neurological disorders tend to be managed within tertiary care settings, necessitating patients needing to travel to specialised medical centres for multidisciplinary care. Monitoring of neuropathological blood tests would likely require specialist neurological review. While specialist services offer potentially more bespoke care for these complex conditions, frequent clinic visits for blood tests or MRI scans could intensify the burden on the patients and their caregivers. The ability for patients to collect their own blood at home for regular monitoring of such biomarkers would therefore be highly advantageous and desirable.

As we move closer to prevention trials for neurodegenerative diseases (5–7) the implications of treating presymptomatic individuals with neuropathological changes need to be considered for clinical trial design. The targeted demographics for such trials will encompass younger individuals who maintain full functional abilities despite their risk of developing a neurodegenerative disease. These individuals carry their own responsibilities, including childcare and early careers which pose challenges to attending the many visits in clinic required for clinical trial participation. Remote sampling for biomarker assessment offers several key benefits: 1) The convenience of self-sampling at home makes repeated sampling easier on participants, which in turn could reduce dropout in long longitudinal studies or trials; 2) Remote sampling could overcome geographical barriers for individuals in areas where a specialist centre is inaccessible; 3) Remote assessments could effectively address challenges associated with recruitment of ultrarare populations. For example, remote testing for sexually transmitted infections almost doubled uptake among ‘never-testers’ (8). Sexual Health London (SHL) and several commercial companies actively provide remote self-sampling services to collect blood via a finger-prick into 600µL microtainer tubes equipped with additives tailored to the post-processing requirements of the desired analytes. Examples include lithium heparin (LiHep) tubes for plasma and serum separator tubes (SST) for serum.

A common blood-based biomarker implicated in many neurological disorders is neurofilament light protein (NfL), a marker of ongoing neuronal damage, which can be measured in both cerebrospinal fluid (CSF) and blood (9). Natural history studies have demonstrated elevation in comparison to age-matched healthy controls in Huntington‘s Disease (HD) (10–15), multiple sclerosis (MS) (16–19), amyotrophic lateral sclerosis (ALS) (20–22), frontotemporal dementia (23), Alzheimer’s disease (24), and its ability to detect changes in those in the very early stages of disease (22, 25, 26). Plasma NfL levels have also shown evidence of being able to distinguish idiopathic Parkinson’s disease (PD) from atypical Parkinsonian syndromes (27–29). NfL also shows potential as a surrogate endpoint with reductions following efficacious treatment demonstrated in MS (30) and spinal muscular atrophy (SMA) (31). Recently, an anti-sense oligonucleotide therapeutic, Tofersen, targeting SOD1 received FDA accelerated approval based on the ability of the drug to lower blood NfL levels in patients with SOD1 mutation-mediated ALS (32).

There have been previous efforts to use small volumes of blood to measure NfL to aid remote monitoring, showing some promise with dried plasma spots (DPS) using Noviplex Plasma Prep cards (33, 34). Several studies have demonstrated that NfL is highly stable in blood plasma and can withstand freeze-thawing and delayed sample processing, even up to seven days (35, 36). However, no studies have empirically tested whether NfL concentrations measured in capillary plasma or serum align with those observed in the established gold standard of venous plasma or serum.

In this study, we adapted the finger prick method used by SHL and applied it to four different neurological conditions (HD, MS, ALS, PD) to assess the validity of quantifying NfL and other exploratory markers of neuronal injury and inflammation via remote sample collection. We collected matched samples from venepuncture and finger-prick and processed them into plasma and/or serum to directly compare NfL levels across a range of control and disease concentrations. We compared two delayed processing conditions, three- and seven-day delay, and simulated ambient shipment. We report NfL concentration from capillary plasma and serum and evidence to support its application for at-home testing.

## Results

### A novel approach to collecting blood for remote NfL quantification

By optimising previously used finger-prick collection methods, we were consistently able to collect two microtainer tubes of capillary blood (600µL each) from each individual finger-prick session. For this reason, each condition comparison needed to be assessed in independent collections. The study design and experimental objectives are outlined in Figure 1. Initially, we set out to compare our novel finger-prick method with previously reported alternative methods for NfL quantification, including dried blood spots (DBS) and DPS (34, 37). However, two pilot collections indicated that the finger-prick collected capillary plasma and serum produced NfL concentrations more similar to the venous blood gold standard than previously published alternative methods (Supplementary Figure 1). Hence, we went on to simply compare our method of finger-prick plasma and serum collection only for NfL analysis to that from the venous blood gold standard. The demographics of each collection are summarised in the supplementary material (Supplementary Table 1). We quantified total protein and haemoglobin for quality control assessment of each collection and sample type. Glial fibrillary acidic protein (GFAP) and total tau (tTau) were quantified as exploratory markers of interest. Summary statistics of all analytes for each experiment are presented in the supplementary material (Supplementary Tables 4-8).

**Figure 1:**
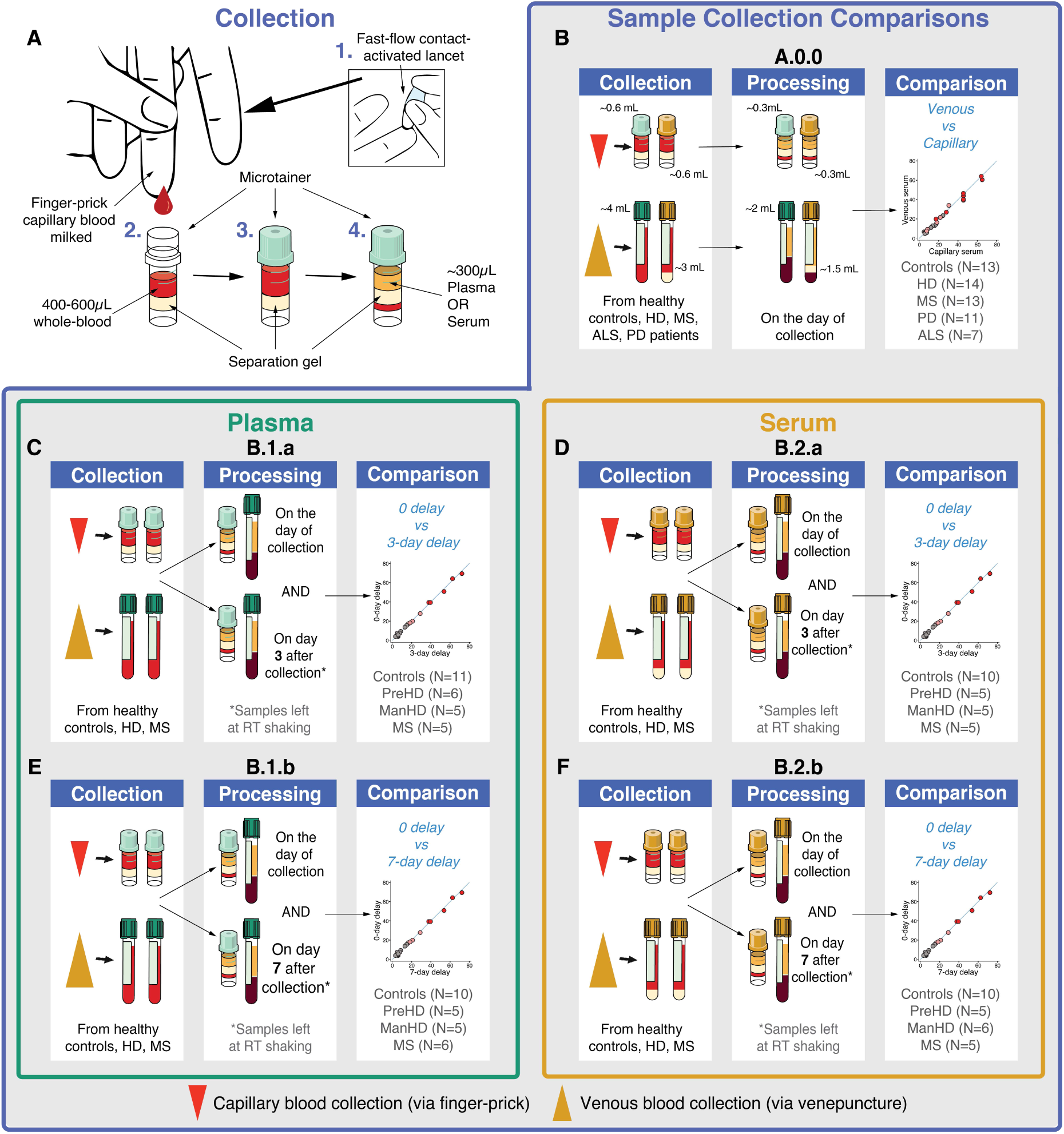
Study design and experimental objectives. A) A schematic showing the capillary sample collection method: 1. Finger-prick performed using a fast-flow lancet, 2. Blood milked from finger, 3. Up to 600µl whole blood collected into plasma (LiHep) or serum (SST) microtainers (2 x objective), 4. ∼300µl capillary plasma/serum generated after processing. B) Experimental objective A.0.0: matched capillary and venous samples were collected in one serum (SST) and one plasma (LiHep) microtainer/vacutainer tube and processed on the day of collection to compare analytes between sample types (plasma/serum) and collection types (capillary/venous). C) Experimental objective B.1.a: matched capillary and venous samples were collected in two plasma microtainer and two plasma vacutainer tubes, one of each collection type was processed on day zero and the other on day three to compare the impact of a three-day delay in processing on plasma analytes. D) Experimental objective B.2.a: matched capillary and venous samples were collected in two serum microtainer and two serum vacutainer tubes, one of each collection type was processed on day zero and the other on day three to compare the impact of a three-day delay in processing on serum analytes. E) Experimental objective B.1.b: matched capillary and venous samples were collected in two plasma microtainer and two plasma vacutainer tubes, one of each collection type was processed on day zero and the other on day seven to compare the impact of a seven-day delay in processing on plasma analytes. F) Experimental objective B.2.b: matched capillary and venous samples were collected in two serum microtainer and two serum vacutainer tubes, one of each collection type was processed on day zero and the other on day seven to compare the impact of a seven-day delay in processing on serum analytes. RT, room temperature; HD, Huntington’s disease; PD, Parkinson’s disease; MS, multiple sclerosis; ALS, amyotrophic lateral sclerosis.

### NfL and GFAP concentrations are the same in venous and capillary plasma and serum

Total protein and haemoglobin concentrations in capillary and venous blood were highly variable across collection and sample types (Figure 2A-J). These assays had similarly low technical variability within runs as those quantified by the ultrasensitive SIMOA method (Supplementary Table 9). There were higher concentrations of total protein in venous blood compared to capillary (Plasma: mean difference (MD) = 9.1 mg/mL, p < 0.0001, Supplementary Figure 2A; Serum: MD = 10.1 mg/mL, p < 0.0001, Supplementary Figure 2B). Despite this, NfL concentrations were strongly correlated between venous and capillary samples in both plasma (r = 0.967, R^2^ = 0.935, p < 0.0001; Figure 2K) and serum (r = 0.963, R^2^ = 0.929, p < 0.0001; Figure 2L), and plasma and serum NfL values were equivalent in both collection types (Venous: r = 0.993, R^2^ = 0.985, p < 0.0001; Figure 2M; Capillary: r = 0.979, R^2^ = 0.957, p < 0.0001; Figure 2N). This was similar for GFAP concentrations (Plasma: r = 0.979, R^2^ = 0.959, p < 0.0001; Serum: r = 0.967, R^2^ = 0.934, p < 0.0001; Venous: r = 0.978, R^2^ = 0.985, p < 0.0001; Capillary: r = 0.973, R^2^ = 0.957, p < 0.0001; Figure 2P-S, respectively). NfL and GFAP values across all four sample and collection types compared in experiment A.0.0 showed strong agreement (NfL: ICC = 0.970, p < 0.0001, Figure 2O; GFAP: ICC = 0.973, p < 0.0001, Figure 2T) and variance did not change with concentration (Bland-Altman limit of agreement < 1 SD of the NfL and GFAP mean differences between different collection types indicating good agreement; Supplementary Figure 3I-P). tTau varied more across sample and collection types (ICC = 0.510, p <0.0001, Figure 2Y), and concentrations were higher in capillary blood compared to venous blood and in plasma compared to serum. Using a post-hoc paired t-test, we found these increases were significant (Plasma: MD = 13.9 pg/mL, p < 0.0001, Supplementary Figure 2E; Serum: MD = 4.7 pg/mL, p < 0.0001, Supplementary Figure 2F) and were higher in plasma samples compared to serum samples in both collection types (Capillary: MD = 13.2 pg/mL, p < 0.0001, Supplementary Figure 2G; Venous: MD = 4.7 pg/mL, p < 0.0001, Supplementary Figure 2H).

**Figure 2:**
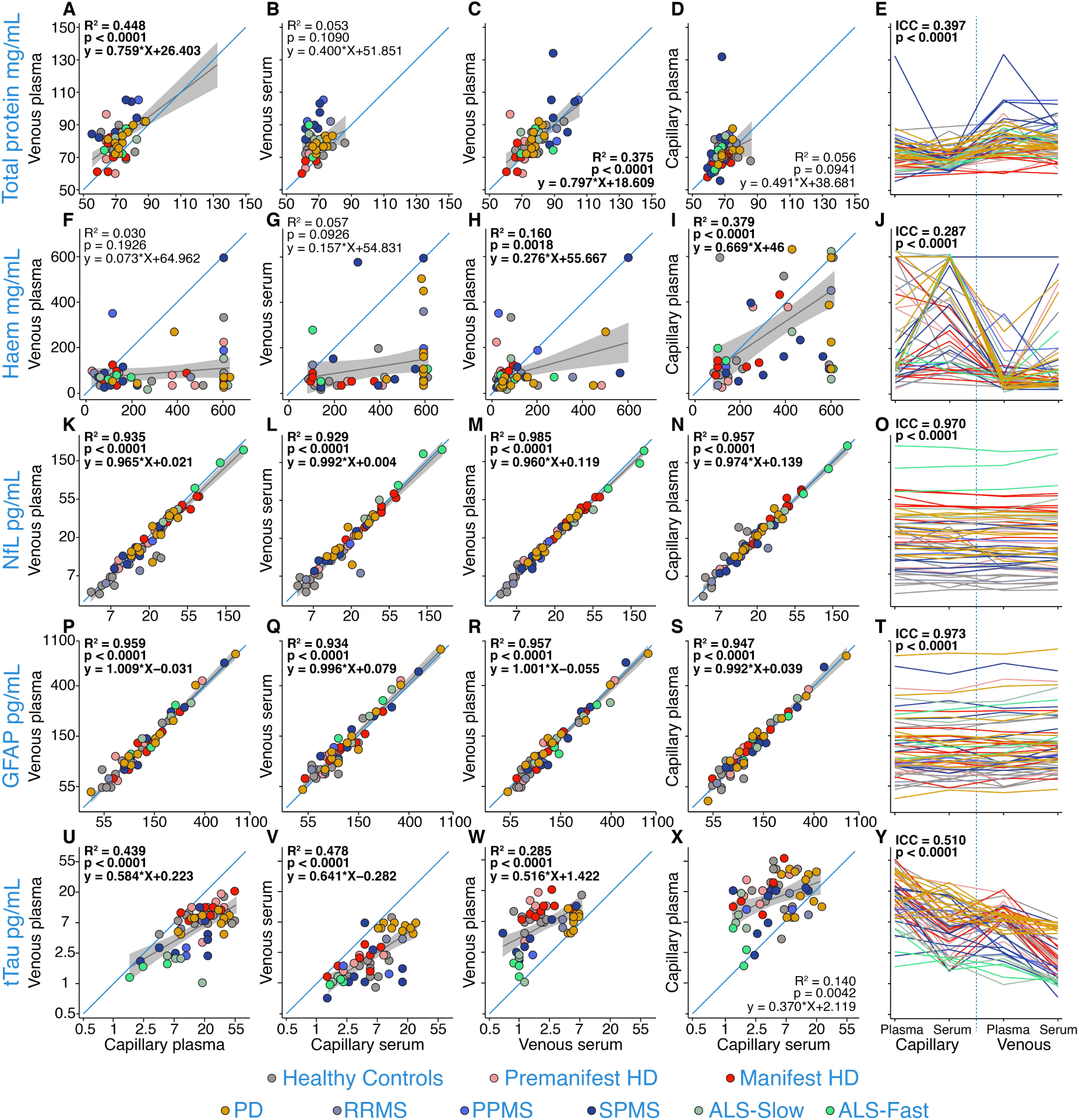
Analyte concentrations across different collection methods and sample types from experiment A.0.0 for total protein (A-E), haemoglobin (F-J), NfL (K-O), GFAP (P-T), and tTau (U-Y) in healthy controls, pre-HD, manifest-HD, PD, RRMS, PPMS, SPMS, ALS-slow, ALS-fast. Blue lines represent y=x. Grey lines represent the linear regression fit of the data with 95% confidence interval shaded regions. R^2^ and p-values were generated from regression models comparing the two collection types in each panel. The Bonferroni threshold for this experiment was 0.0025 (20 comparisons) and all statistics which reached significance below this are highlighted in bold. NfL, GFAP, and tTau concentrations were natural log-transformed. HD, Huntington’s disease; PD, Parkinson’s disease; RRMS, relapsing-remitting multiple sclerosis; PPMS, primary progressive multiple sclerosis; SPMS, secondary progressive multiple sclerosis; ALS, amyotrophic lateral sclerosis; Haem, haemoglobin; tTau, total tau.

### NfL shows high stability after three-day delayed processing in venous and capillary plasma and serum

After confirming capillary blood NfL was equivalent to venous blood NfL, we tested the impact of a three-day delay in processing on analyte concentrations in plasma (B.1.a) and serum (B.2.a) from healthy controls, HD and MS participants. There were clear trends of increase in both total protein and haemoglobin concentrations in venous and capillary plasma after a three-day delay in processing (Capillary total protein: Figure 3A-E; MD = 9.19 mg/mL, p < 0.0001, Supplementary Figure 4A; Capillary haemoglobin: Figure 3F-J; MD = 135.5 mg/mL, p = 0.0008, Supplementary Figure 4C; Venous haemoglobin: Figure 3F-J; MD = 245.6 mg/mL, p < 0.0001, Supplementary Figure 4D). In serum, only venous haemoglobin showed signs of elevation (Figure 4F-J; MD = 240.1 mg/mL, p < 0.0001, Supplementary Figure 5B) after a three-day delay in processing.

**Figure 3:**
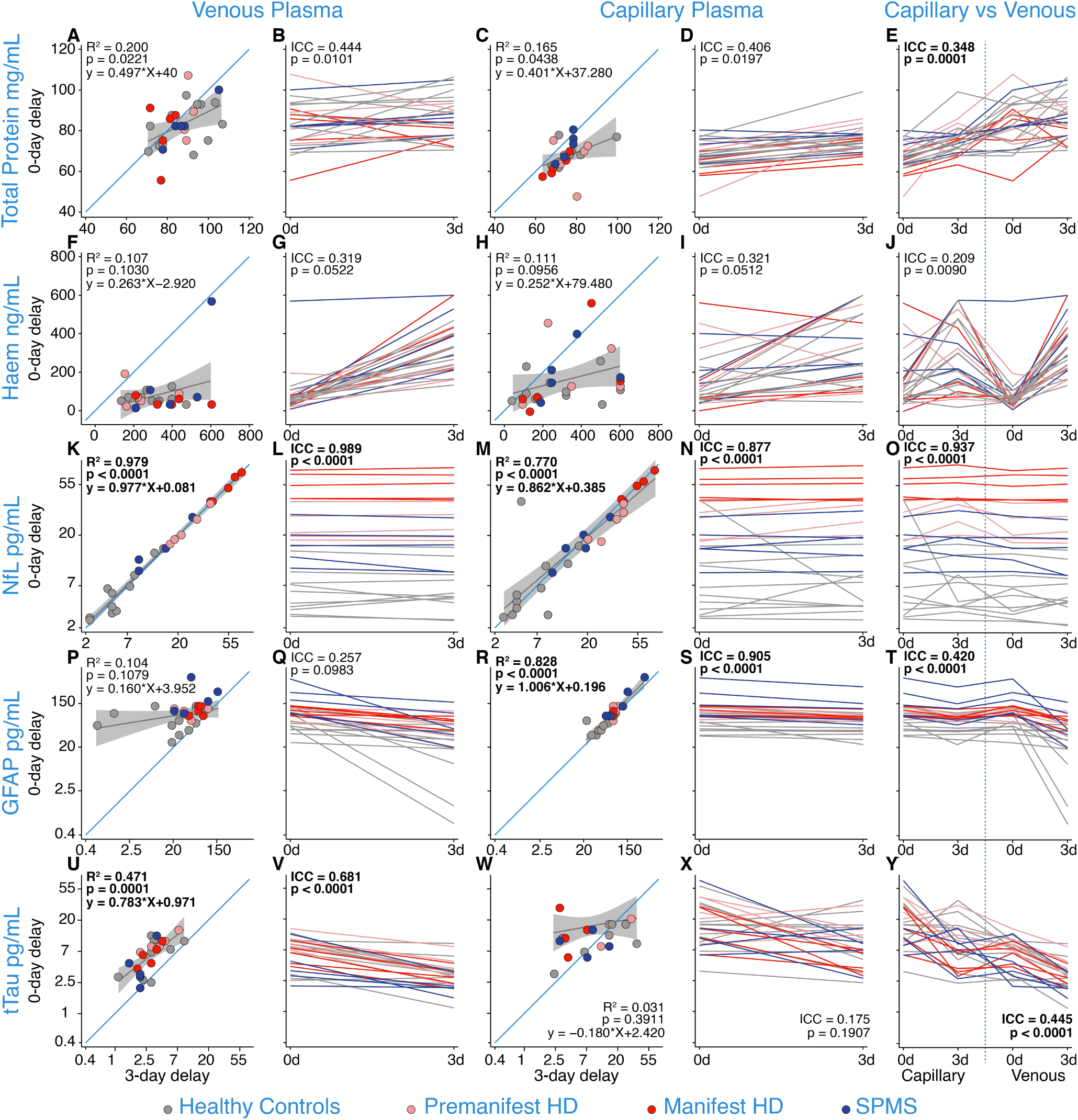
Analyte concentrations in plasma after zero-day and three-day delay in processing from experiment B.1.a. for total protein (A-E), haemoglobin (F-J), NfL (K-O), GFAP (P-T), and tTau (U-Y) from venous and capillary collections in healthy controls, pre-HD, manifest-HD, SPMS, Blue lines represent y=x. Grey lines represent the linear regression fit of the data with 95% confidence interval shaded regions. R^2^ and p-values were generated from regression models comparing the impact of delayed processing in each panel. The Bonferroni threshold for this experiment was 0.005 (10 comparisons) and all statistics which reached significance below this are highlighted in bold. NfL, GFAP, and tTau concentrations were natural log-transformed. HD, Huntington’s disease; SPMS, secondary progressive multiple sclerosis; Haem, haemoglobin; tTau, total tau.

**Figure 4:**
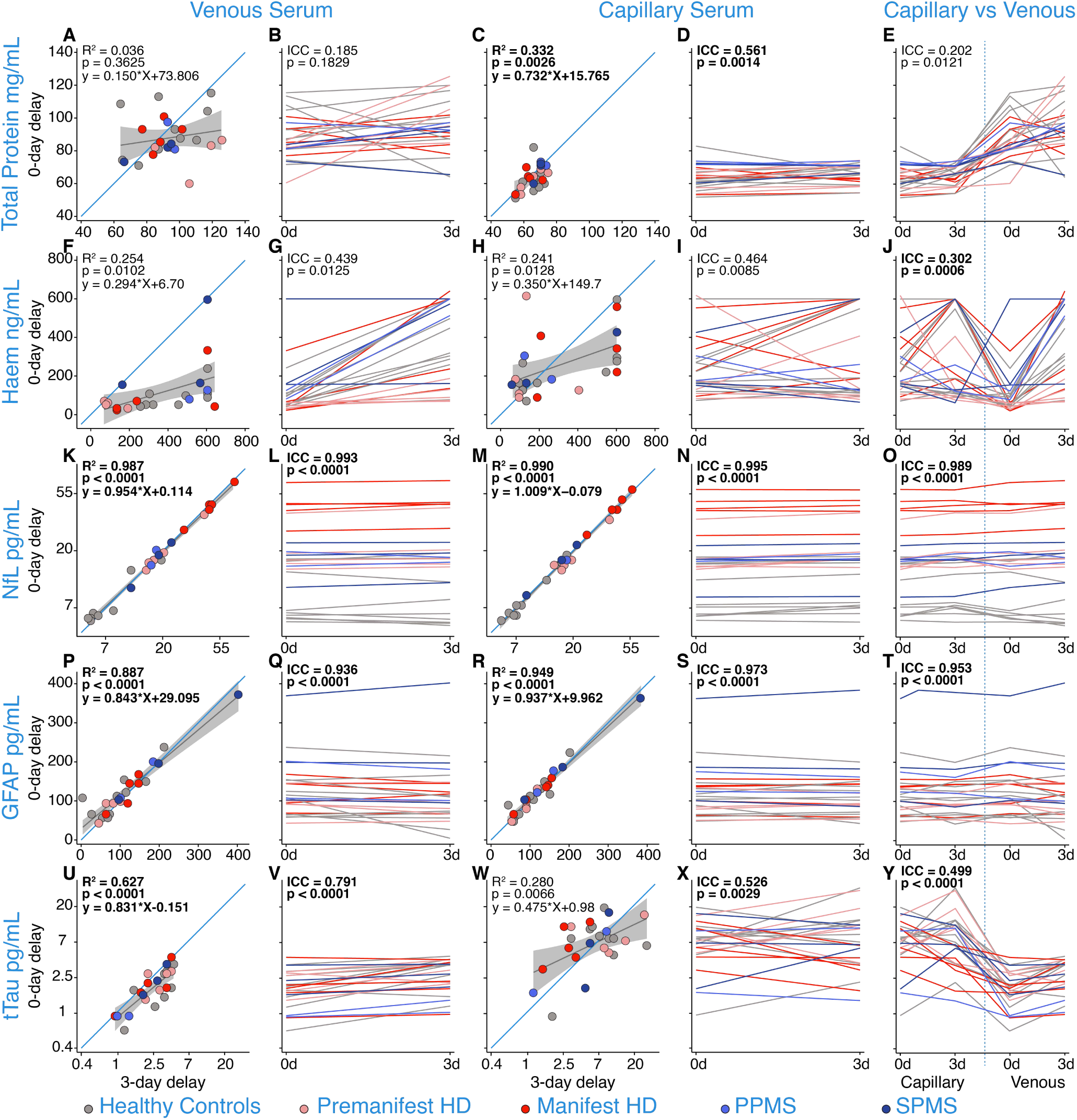
Analyte concentrations in serum after zero-day and three-day delay in processing from experiment B.2.a. for total protein (A-E), haemoglobin (F-J), NfL (K-O), GFAP (P-T), and tTau (U-Y) from venous and capillary serum in healthy controls, pre-HD, manifest-HD, PPMS, SPMS. Blue lines represent y=x. Grey lines represent the linear regression fit of the data with 95% confidence interval shaded regions. R^2^ and p-values were generated from regression models comparing the impact of delayed processing in each panel. The Bonferroni threshold for this experiment was 0.005 (10 comparisons) and all statistics which reached significance below this are highlighted in bold. NfL and tTau concentrations were natural log-transformed. HD; Huntington’s disease; PPMS, primary progressive multiple sclerosis; SPMS, secondary progressive multiple sclerosis; Haem, haemoglobin; tTau, total tau.

Plasma and serum NfL concentrations showed strong positive correlations between samples processed on the day of collection versus three days later for both venous (Plasma: r = 0.989, R^2^ = 0.979 p < 0.0001, Figure 3K-L; Serum: r = 0.993, R^2^ = 0.987 p < 0.0001, Figure 4K-L) and capillary (Plasma: r = 0.877, R^2^ = 0.770 p < 0.0001, Figure 3M-N; Serum: r = 0.995, R^2^ = 0.990 p < 0.0001, Figure 4M-N) samples, suggesting high stability of NfL after three days of delayed processing, irrespective of collection method. The Bland-Altman analysis confirmed strong agreement across the NfL concentrations measured in both serum and plasma between zero- and three-day samples (Supplementary Figure 9I-L). Similarly, GFAP concentrations showed strong positive correlations in capillary plasma (r = 0.919, R^2^ = 0.828, p < 0.0001, Figure 3R-S) and serum (r = 0.929, R^2^ = 0.949, p < 0.0001, Figure 4R-S) after three days delay in processing compared to day zero concentrations. However, GFAP in venous plasma was more variable than in capillary plasma after three days delay (R^2^ = 0.104 and R^2^ = 0.828, respectively; Figure 3P&R); in venous plasma, GFAP concentrations were significantly reduced after a three-day delay (Figure 3P-Q; MD = 70.8 pg/mL, p = 0.0001, Supplementary Figure 4F) and in capillary plasma (Figure 3R-S; MD = 32.4 pg/mL, p = 0.0001, Supplementary Figure 4E). There were no significant differences between serum capillary or venous GFAP concentrations between processing on day zero and day three (Capillary: Figure 4R-S, MD = 0.5 pg/mL, p = 0.8629, Supplementary Figure 5C; Venous: Figure 4P-Q, MD = 10.1 pg/mL, p = 0.0976, Supplementary Figure 5D). Capillary tTau concentrations with three-day delayed processing were again highly variable in plasma (Figure 3X, ICC = 0.175, p = 0.1907) and serum (Figure 4X, ICC = 0.526, p = 0.0029) samples. There was a marked decrease in capillary plasma tTau after three-day delayed processing (Figure 3X; MD = 10.8 pg/mL p = 0.0072, Supplementary Figure 4G) also reflected in venous plasma (Figure 3V; MD = 4.3 pg/mL, p < 0.0005, Supplementary Figure 4H).

### NfL shows high stability after seven-day delayed processing in venous and capillary plasma and serum

After assessing the stability of NfL following a three-day delay in processing, we proceeded to extend this delay and examine the impact of a seven-day processing delay on analyte concentrations in Plasma (B.1.b) and Serum (B.2.b) from healthy controls, HD, and MS participants. Total protein concentrations remained highly variable with seven days of delayed processing in plasma (Venous: r = 0.166, R^2^ = 0.028, p = 0.4167, Figure 5A-B; Capillary: r = 0.294, R^2^ = 0.087, p = 0.1535, Figure 5C-D) and serum (Venous: r = 0.389, R^2^ = 0.151, p = 0.0495, Figure 6A-B; Capillary: r = 0740, R^2^ = 0.548, p < 0.0001, Figure 6C-D). Haemoglobin concentrations dramatically increased in samples processed with a seven-day delay compared to those processed on the day of collection for both venous (Plasma: Figure 5F-G; MD = 5.3 ug/ml, p < 0.0001, Supplementary Figure 6B; Serum: Figure 6F-G; MD = 5.1 ug/ml, p < 0.0001, Supplementary Figure 7B) and capillary (Plasma: Figure 5H-I; MD = 4.4 ug/ml, p < 0.0001, Supplementary Figure 6A; Serum: Figure 6H-I; MD = 4.4 ug/ml, p < 0.0001, Supplementary Figure 7A) samples.

**Figure 5:**
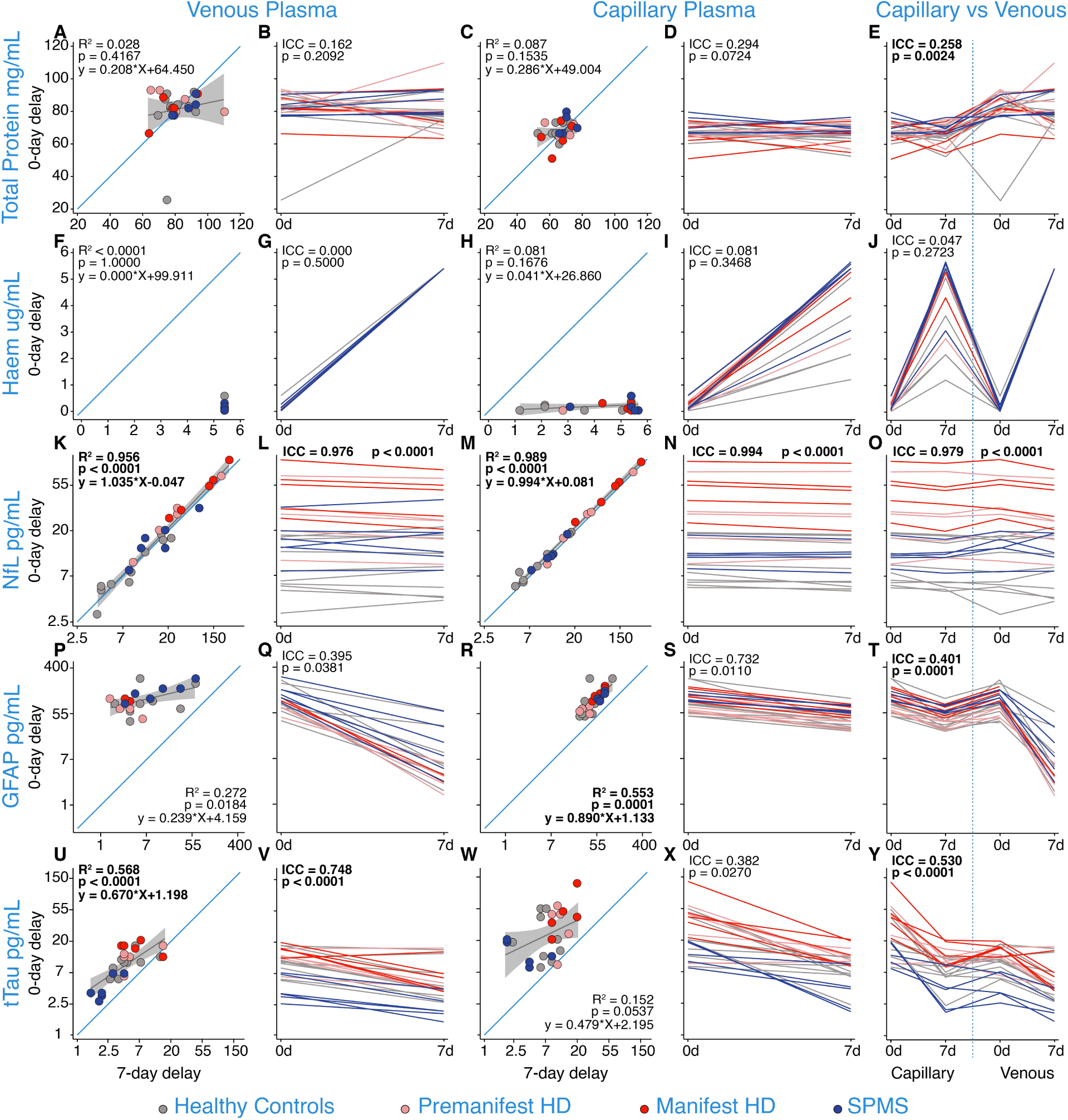
Analyte concentrations in plasma after zero-day and seven-day delay in processing from experiment B.1.b. for total protein (A-E), haemoglobin (F-J), NfL (K-O), GFAP (P-T), and tTau (U-Y) from venous and capillary plasma in healthy controls, pre-HD, manifest-HD, and SPMS. Blue lines represent y=x. Grey lines represent the linear regression fit of the data with 95% confidence interval shaded regions. R^2^ and p-values were generated from regression models comparing the impact of delayed processing in each panel. The Bonferroni threshold for this experiment was 0.005 (10 comparisons) and all statistics which reached significance below this are highlighted in bold. NfL, GFAP, and tTau concentrations were natural log-transformed. HD; Huntington’s disease; SPMS, secondary progressive multiple sclerosis; Haem, haemoglobin; tTau, total tau.

**Figure 6:**
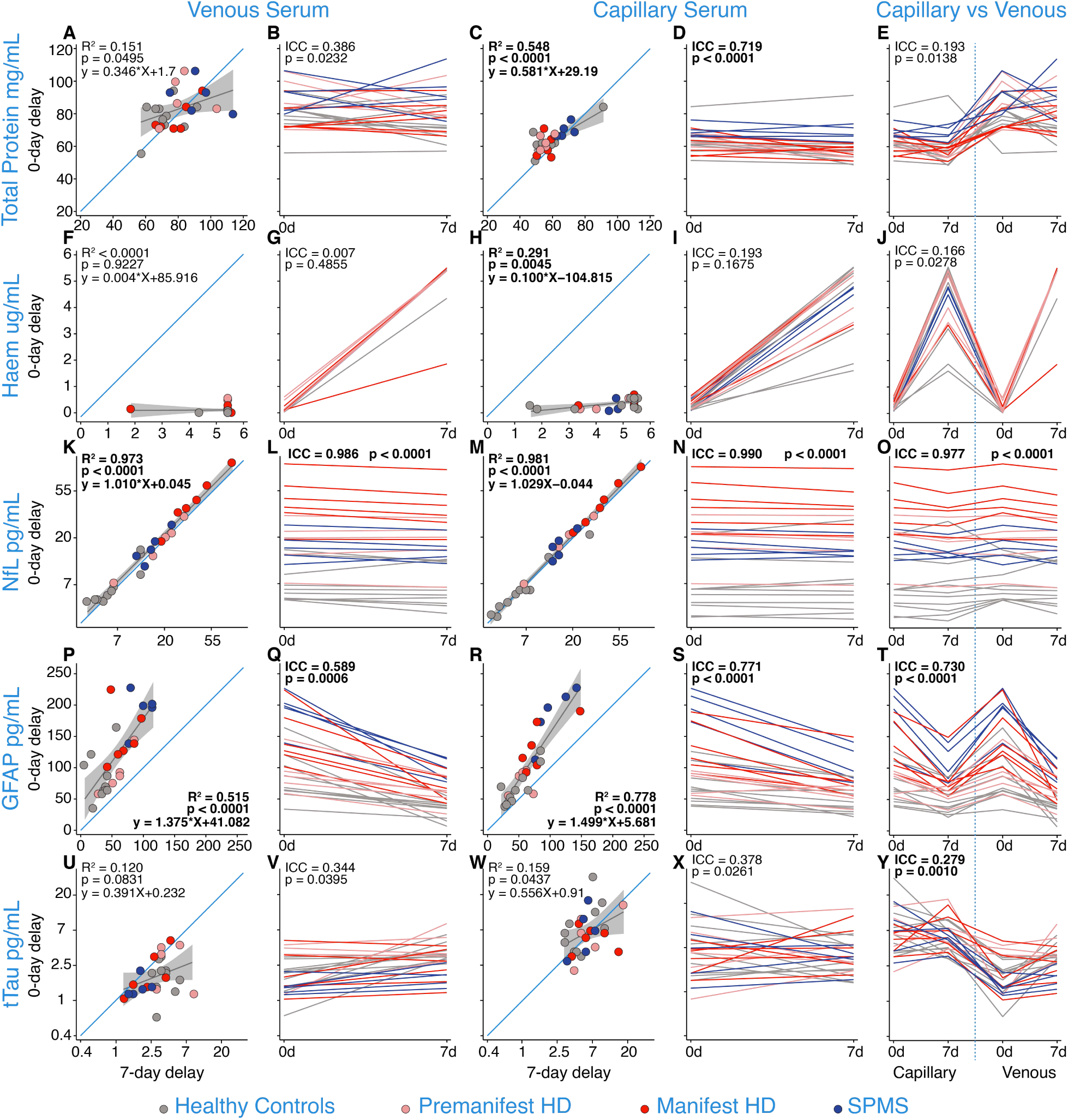
Analyte concentrations in serum after zero-day and seven-day delay in processing from experiment B.2.b for total protein (A-E), haemoglobin (F-J), NfL (K-O), GFAP (P-T), and tTau (U-Y) from venous and capillary serum in healthy controls, pre-HD, manifest-HD, and SPMS. Blue lines represent y=x. Grey lines represent the linear regression fit of the data with 95% confidence interval shaded regions. R^2^ and p-values were generated from regression models comparing the impact of delayed processing in each panel. The Bonferroni threshold for this experiment was 0.005 (10 comparisons) and all statistics which reached significance below this are highlighted in bold. NfL and tTau concentrations were natural log-transformed. HD; Huntington’s disease; SPMS, secondary progressive multiple sclerosis; Haem, haemoglobin; tTau, total tau.

Plasma and serum NfL concentrations show robust positive correlations when samples were processed on the day of collection versus seven days later for both venous (Plasma: r = 0.978, R^2^ = 0.956 p < 0.0001, Figure 5K-L; Serum: r = 0.986, R^2^ = 0.973 p < 0.0001, Figure 6K-L) and capillary (Plasma: r = 0.994, R^2^ = 0.989 p < 0.0001, Figure 5M-N; Serum: r = 0.991, R^2^ = 0.981 p < 0.0001, Figure 6M-N) samples. The Bland-Altman analysis confirmed strong agreement across the NfL concentrations measured in both serum and plasma between zero- and seven-day delay processed samples (Supplementary Figure 10I-L). This stability was not reflected in any of the other analytes. GFAP concentrations decline with seven days of delayed processing in both venous (Plasma: Figure 5Q, MD = 101.1 pg/mL, p < 0.0001, Supplementary Figure 6D; Serum: Figure 6Q, MD = 62.8 pg/mL, p < 0.0001, Supplementary Figure 7D) and capillary (Plasma: Figure 5S, MD = 56.9 pg/mL, p < 0.0001, Supplementary Figure 6C; Serum: Figure 6S, MD = 39.9 pg/mL, p < 0.0001, Supplementary Figure 7C) samples. Concentrations of plasma tTau declined in both capillary (Figure 5X; MD = 22.4 pg/mL, p < 0.0001, Supplementary 6E) and venous (Figure 6V; MD = 4.9 pg/mL, p < 0.0001, Supplementary 6F) samples after seven days delay in processing and remain highly variable in serum venous (r = 0.346, R^2^ = 0.120, p = 0.0831, Figure 6U-V) and capillary (r = 0.399, R^2^ = 0.159, p = 0.0437, Figure 6W-X) samples.

### Capillary NfL shows the same clinical disease group differences as venous NfL

As an exploratory analysis, we combined samples processed on the day of collection from all experiments to compare NfL concentrations between clinical disease groups and healthy controls for each sample and collection type (Figure 7). The same pattern of disease group differences was seen for all collection and sample types. Compared to controls, pre-manifest HD (pre-HD), manifest-HD, and ALS groups showed significantly elevated NfL concentrations for all collection types when controlling for age with similar MD (all p-values < 0.0001, Figure 7A-D, Supplementary Table 2). For each collection type, MS groups showed no significant difference in NfL concentrations compared to healthy controls apart from in capillary serum samples which showed significantly higher NfL levels but did not survive Bonferroni correction (p = 0.027, Figure 7D) and MD was similar (Supplementary Table 2). PD had significantly elevated NfL levels from healthy controls in venous plasma, capillary plasma and serum samples but only capillary serum survived Bonferroni correction (MD = 12.1 pg/mL, p = 0.010, Figure 7A; MD = 11.2 pg/mL, p = 0.024, Figure 7C; MD = 11.8, p = 0.0023, Figure 7D). There were no significant differences between disease groups and healthy controls for GFAP measures and this was consistent across sample and collection types (Supplementary Figure 8, Supplementary Table 3).

**Figure 7:**
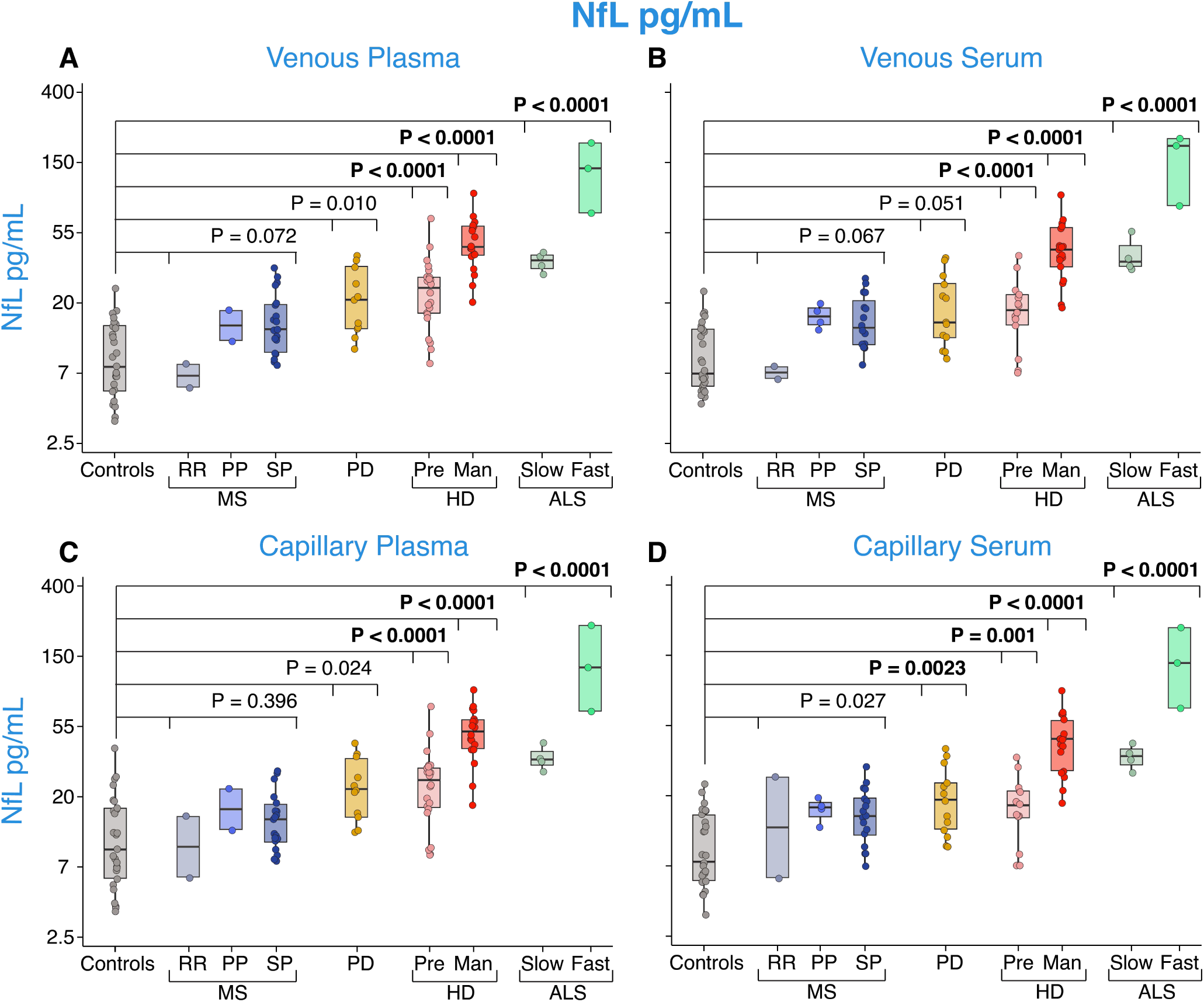
Disease group comparison of NfL concentrations from venous plasma (A), venous serum (B), capillary plasma (C), and capillary serum (D) from all baseline data for healthy controls, PPMS, RRMS, SPMS, PD Pre-HD, manifest-HD, ALS-slow and ALS-fast patients. p-values were generated from multiple linear regressions. The Bonferroni threshold for this experiment was 0.0025 (20 comparisons) and all statistics which reached significance below this are highlighted in bold. NfL values are natural log-transformed. PD, Parkinson’s disease; RR, relapsing-remitting; PP, primary progressive; SP, secondary progressive; MS, multiple sclerosis; HD, Huntington’s disease; Pre, premanifest; Man, manifest; ALS, amyotrophic lateral sclerosis.

## Discussion

In this study, we investigated the potential of a novel finger-prick blood collection method for remote quantification of NfL in four different neurological conditions (HD, MS, ALS, PD) via a series of experiments that directly compared the impact of the collection method, processed sample type, and delayed processing on analyte concentrations. Our findings demonstrate that NfL and GFAP concentrations were equivalent whether measured from capillary or venous blood collections and whether processed into serum or plasma. NfL was able to withstand delayed processing of seven days.

The methodologies employed in this study represent, to the best of our knowledge, the first use of capillary finger-prick blood collection techniques for biomarker assessment in neurological disorders. Previous reports of alternative collection and processing methods for NfL quantification, such as DPS using Noviplex Plasma Prep cards, were performed using venous blood, and despite being highly correlated, DPS NfL values were approximately four times lower than standard venepuncture and processing NfL concentrations (33, 34). Here, we report regression fits and data points for each condition comparison of NfL concentrations to lie on the y=x line, an indication of equivalence.

Despite variations in haemolysis produced by each method, concentrations of NfL and GFAP remained consistent in both venous and capillary samples irrespective of processed sample type; this mirrors previous studies that have investigated the translatability of NfL and GFAP between serum and plasma samples (38). This marks an advancement in our understanding of the feasibility and accuracy of this minimally invasive approach to quantify a key biomarker of neuronal injury. That GFAP concentrations also demonstrate strong agreement between venous and capillary blood is promising, as it supports the development of additional blood-based biomarkers for remote quantification.

The robust agreement between sample and collection type for NfL and GFAP was not replicated in other analytes: there was high variability in tTau levels between capillary and venous blood, as well as between serum and plasma in each collection method. This is in line with previous reports that found no overall correlation between plasma and serum tTau levels (38, 39). Consequently, considerations regarding the choice of collection method and sample type (venepuncture, capillary, serum, or plasma) need to be further assessed for blood tTau measurements. Furthermore, tTau displayed considerable variability between sample types with delayed processing, suggesting tTau in any sample type or collection method is not suitable for remote collections.

Our results demonstrate that NfL exhibits stability up to seven days of delayed processing in both venous and capillary samples. This replicates the stability of NfL in venous blood presented in previous studies (40–43) and further shows its stability in capillary blood samples. In contrast, GFAP concentrations declined after a seven-day delay in processing with signs of reduction even at three-day delay. Given the persistence of strong correlations despite these reductions in GFAP concentrations, and the stepwise decreases with delay interval, the impact of delayed processing on GFAP may be consistent and quantifiable. If this can be quantified, it could support the use of a conversion factor to account for the impact of delayed processing. This is outside the scope of the current study but may warrant further study for disease conditions where GFAP is a strong candidate biomarker.

The delay interval investigated in this study spanned seven days; the longest delay interval studied for venous blood NfL is eight days (41). The potential of a remote sample collection for NfL quantification could facilitate a wide-scale international study of this biomarker. The complexities associated with international shipping increase the risk of longer processing delays where samples may take more than a week for delivery. Therefore, further studies should consider investigations extending beyond this duration in capillary samples.

The impact of delayed processing on analyte concentrations differs in dynamics for all analytes. By quantifying total protein and haemoglobin, we could begin to assess the generalised impact of collection, sample type, and processing delay on proteins in the blood that could present confounding effects on biomarker quantification. As expected, haemoglobin increased with increasing delay in sample processing, consistent with the increased haemolysis visibly seen when processing these samples. The subtle increases in total protein after three days and lack of change after seven days of delayed processing could be due to the increased haemolysis causing subtle increases in total protein concentrations outweighing any protein degradation (44, 45). We would expect haemolysis to reach a maximum with further increases in delay interval and for total protein to begin to decrease as proteins begin to degrade. This needs to be investigated further.

Both NfL and GFAP have broad and important relevance across multiple neurological disorders as blood-based biomarkers. Although this study aimed to technically validate our finger-prick collection method for remote quantification of NfL, combining data from all experiments permitted us to assess capillary blood NfL to replicate previously reported disease group differences. We have reproduced previous findings that NfL is significantly elevated in HD (10–12), PD (27, 46) and ALS (22, 47) compared to controls. The clear NfL profile in these cohorts suggests that our remote sampling method for NfL measurement may be useful in these disease populations and potentially in other neurological disorders.

NfL could have several applications in therapeutic development and clinical management in neurology. NfL has been proposed as a potential surrogate endpoint indicating neuroprotection for multiple neurological disorders (e.g., MS (48–50), ALS (51–53), and HD (10, 54, 55)); its utility as a safety biomarker for neurotoxicity is also being considered (56, 57); and in the future NfL might be used for enrichment and stratification of presymptomatic populations to facilitate the design of prevention trials (10, 54, 55). However, before NfL measurements can be validated for either intended use, there needs to be deep characterisation of its natural fluctuations throughout the progression of each disease. Remote collection offers the potential to facilitate frequent short-interval sampling and real-world monitoring of patients. Not only would this reduce the burden of in-person visits on patients but could also expedite indications of drug efficacy, drug toxicity, or clinical progression. For example, an increase in NfL levels, mirroring increasing rates of neuronal injury, could indicate that a disease-modifying treatment may need to be initiated. NfL increases in direct response to the initiation of a new treatment could quickly inform clinicians to discontinue this treatment or change their therapeutic approach. Monitoring NfL levels throughout disease progression may aid preclinical staging and inform clinical care decisions and planning. Large-scale remote collections for frequent sampling and quantification of NfL are now needed to fully characterise blood NfL over the entire natural history of neurological disorders for these applications.

Our study has several limitations: First, accurately simulating blood sample shipment conditions in controlled experiments is extremely difficult. In our study, samples were left for three or seven days on a shaker at room temperature in our laboratory. This did not simulate the significant temperature fluctuations that sample kits may endure whilst being shipped from the patients’ homes to the laboratory. The temperature that parcels are exposed to varies greatly depending on the mode of transportation, weather conditions, duration of transport and specific handling procedures from the courier service. A study investigated the temperatures that 72 shipments were exposed to and showed that although the average temperature the packages were exposed to was 26.2 ± 2.3°C (near room temperature), they frequently sustain temperature spikes above 40°C (highest recorded at 52.9°C for over 12 hours) (58). Previous data has shown that NfL can sustain multiple freeze-thaw cycles (9, 35, 36, 59), and up to 24 hours at 37°C incubation (33, 34). There is no evidence to our knowledge that confirms that it can sustain higher temperatures. Furthermore, a shaker cannot replicate the unpredictable mechanical stresses that the samples go through during shipping. Indeed, samples may fall during transport or could be left still for many hours at a time. It may be important to repeat our experiment leaving the samples still on the bench, inducing random mechanical stresses, increasing delay intervals, and adding varied temperature protocol to determine whether this affects NfL stability.

Second, although we showed finger-prick NfL reproduced similar disease group differences from previously reported cohort studies, our study was not powered for this post-hoc analysis. At-home collections will need to be set up and assessed for each disease aiming to validate remote NfL quantifications for its population. Third, as this was a technical validation of the method and collections were performed by our team, we have not yet shown the feasibility of patients performing self-sampling at home without a health professional guiding them. However, we have already established ongoing remote collections with over 100 individuals (premanifest HD and matched healthy controls) actively collecting blood in their own home and returning the samples by post to our laboratory every two months. We are planning other studies in clinical populations to address this further. Finally, for tTau, the recommended sample matrix is EDTA plasma (60), which was not available in our study. We cannot exclude that remote collection of capillary EDTA plasma would produce better results.

In conclusion, this study has shown that the capillary finger-prick sample collection methods used for NfL measurement may be reliable and useful in several disease populations. According to our results, NfL can be measured from capillary plasma and serum seven days after remote collection, with equivalent concentrations to the current gold standard of plasma and serum collection (in-clinic venepuncture with same-day blood processing and storage). A crucial element in accelerating patient-centred healthcare involves the capability to remotely collect blood samples with the same quality as traditional phlebotomy that facilitate regular monitoring of disease progression and test drug responses to therapeutic intervention. With further validation, we believe this method could be used for remote monitoring of NfL levels in neurodegenerative disease populations, with higher sampling frequency and geographical patient outreach than in-clinic blood collections could accommodate. The applications for NfL across the spectrum of neurological disorders emphasise the widespread translatability of these methods, which have the potential to transform disease monitoring, prognosis, and therapeutic development within clinical trials and practice.

## Methods

### Study design

This study was designed to develop a finger-prick blood collection method and validate it for remote quantification of NfL in four neurological conditions (HD, MS, ALS, and PD). Healthy controls, pre-manifest HD (pre-HD) and manifest-HD participants were recruited from the National Hospital for Neurology and Neurosurgery (NHNN) HD multidisciplinary clinic, part of University College London Hospitals NHS Foundation Trust. ALS participants were recruited from the Lighthouse II study, ALS biomarkers study, and ALS clinic. MS participants were recruited from the UCL Institute of Neurology Clinical trials (MS-STAT2 and Octopus) and NHS outpatients. PD participants were recruited from East London Parkinson’s Disease project, Royal London Hospital and Barts Health NHS Trust. Participants were aged 18 – 84 years, able to tolerate blood collection, and without major psychiatric disorder or history of significant head injury. Clinical information and disease diagnosis were acquired either from the participant’s medical records or from their clinical team.

Five experiments (Figure 1) were designed to compare NfL concentrations from A.0.0) venous versus capillary and serum versus plasma; B.1.a) venous or capillary plasma with a three-day delay in processing; B.1.b) venous or capillary plasma with a seven-day delay in processing; B.2.a) venous or capillary serum with a three-day delay in processing; B.2.b) venous or capillary serum with a seven-day delay in processing. Total protein and haemoglobin were quantified to assess sample quality and the impact of delayed processing on protein degradation and haemolysis, respectively. GFAP and tTau were quantified as exploratory biomarkers for remote quantification. Sample sizes were derived using a one-sample correlation test (alpha = 0.01, power = 0.9) based on observed correlations of NfL between venous blood and CSF (r = 0.7 – 0.9) (13). N=23 had sufficient power to detect a correlation of r = 0.7 or higher. Therefore, for each objective, at least 23 participants were recruited (total N with all disease groups).

### Blood collection and processing

Venous blood was collected via venepuncture using a butterfly needle. We developed a finger-prick blood collection method to apply to NfL quantification (Figure 1). Blood flow was increased by warming the participants hand and getting them to stand with hands below the heart where possible. Capillary whole blood was gently milked and collected in microtainer tubes (400-600µl per tube; BD SST serum gold top [Product code: 365968] or BD PST LiHep plasma green top [Product code: 12957646].

Delayed processing experiments to assess the impact of delayed processing on NfL in both plasma and serum included collecting two tubes of either plasma or serum for both capillary and venous blood collection. One set of capillary and venous tubes was processed immediately on the collection day, while the other set was left on a shaker for either three or seven days before processing to simulate delays from ambient shipment of at-home collected samples.

Blood from both vacutainers and microtainers was processed using the manufacture’s recommended settings on-site to isolate serum or plasma, either on the day of or three or seven days after sample collection. Vacutainer tubes were centrifuged at 1300g, for 10 minutes at room temperature and the supernatant was isolated, aliquoted into 500µL aliquots. Microtainers were centrifuged at 15000g for 2 minutes at room temperature and the supernatant was isolated, aliquoted into 100µL aliquots. Processed samples were then stored at -80°C until analyte quantification.

### Analyte quantification

NfL, GFAP, and tTau concentrations were quantified using a commercially available Neurology 4-plex-B (N4PB) kit on the SIMOA HD-X analyser platform following the manufacturer’s instructions (Quanterix, Billerica, MA). Control and samples were run in duplicates with machine dilution of 1:4. Haemoglobin was measured in duplicates using a commercial ELISA (Bethyl Laboratories, cat#E88-134) according to the manufacturer’s specifications. Samples processed on the day of collection and three days later were diluted 1:3000 and samples that were processed seven days after collection were diluted 1:27000 to keep samples in the linear range of the assay. Total protein was also measured using a commercial ELISA kit according to manufacturer instructions with samples diluted 1:100 (Pierce™ BCA Protein Assay Kits, cat#23227). Total protein and haemoglobin were measured for each sample from the same aliquot as the N4PB analysis.

For all analytes, each sample within an objective was run using the same batch of reagents and samples from the same individual were run on the same plate. The inter-assay Coefficients of Variance (CV) and average intra-assay CVs for each analyte are represented in Supplementary Table 9. All analyte concentrations fell within the linear range of each assay. Quantification of analytes was performed blinded to disease status.

### Statistical analysis

Analyses were performed using Stata/MP 18.0. The significance level was defined as p < 0.05. Outliers were assessed using box plots and spike plots of the raw data; one outlier was identified with NfL concentrations more than eight times the second highest value (77pg/mL to 653pg/mL) and a GFAP value more than 26 times the second highest value (290pg/mL to 7789pg/mL). Therefore, this participant was excluded from all further analyses. Whole group demographics and group demographics by experiment for age were compared using the Kruskal-Wallis test as data were non-normally distributed. Sex group differences were assessed using Pearson’s X^2^ test.

Normality was assessed visually and using Shapiro-Wilk tests for each comparison grouping in all experiments. NfL and tTau were non-normally distributed for each sample collection type, so natural log-transformed values were used for all experiments. GFAP had non-normal distributions for groups in experiments A.0.0, B.1.a, and B.1.b which was resolved by natural log transforming the data for these experiments.

Assessment of relationships between sample types and/or delayed processing regimens on analyte concentration for each objective was assessed using linear regression models (R^2^) and Pearson’s correlations (r). All p-values were adjusted for multiple comparisons by defining Bonferroni corrected thresholds for each experiment. Two-way mixed-effects model intraclass correlation coefficients (ICC) were used to measure the agreement of analyte measurements from all sample types in each objective with values ranging from zero to one. Bland-Altman plots were also created to visualise the agreement between measurements by analyte concentrations for each objective (Supplementary Figures 3, 9, 10). Where there was visually an increase or decrease between analyte values across sample types/conditions, we performed post-hoc two-tailed paired t-tests to assess whether the difference was significant. To explore group analyte differences between clinical disease groups and controls, we used all baseline samples across all objectives for each sample type: day zero concentrations for venous and capillary plasma and serum. Only NfL and GFAP were assessed for inter-group differences due to their high correlations between sample and collection types. Natural log transformations of NfL and GFAP were used due to non-normally distributed data across sample types and groups. Potentially confounding demographic variables (age, sex) were examined in preliminary analyses; age was identified as a confounding variable for disease group difference and was therefore included as a covariate in subsequent analyses. Multiple linear regression with post-estimation Wald tests was used to assess intergroup analyte concentrations for each sample type with Bonferroni correction for multiple comparisons.

## Study approval

This study complies with the Declaration of Helsinki and is approved by local ethics committees, and all participants gave written informed consent before blood collection. Data and sample collections for this study had ethical approval from The National Hospital for Neurology and Neurosurgery and the Institute of Neurology Joint Research Ethics Committee (HD and controls; ref: 03/N008); London - City & East Research Ethics committee (ALS, MS; ref: 09/H0703/27); South West – Central Bristol Research Ethics Committee (PD: PR 18/SW/0255).

## Data Availability

All data produced in the present study are available upon reasonable request to the authors

## Author contributions

Designing research studies: LB

Recruitment: LB AC AT MF MN MJM YA OT AA ARGC HS LZ VL EC KD AZ SF MN KF CW BF SM

Sample collection: LB AC AT MF MJM OT SF KF EC KCD AZ AA ARGC HS LZ BF

Sample processing: LB AC AT MF MP OT SF EB BH ND AA KF

Analyte quantification: LB AC AT MF

Data analysis: LB AC AT

Writing manuscript: LB AC AT

Reviewing manuscript: AC AT MF MP MJM YH OT SF MN EB BH ND EC KD AZ AA ARGC HS LZ VL KF CW BF SM AH HZ AN AM JC SJT LBl

## Acknowledgments

This work and the salaries of LMB, AC, and AT were supported by a Medical Research Council Career Development Award (MR/W026686/1). HZ is a Wallenberg Scholar supported by grants from the Swedish Research Council (#2022-01018 and #2019-02397), the European Union’s Horizon Europe research and innovation programme under grant agreement No 101053962, Swedish State Support for Clinical Research (#ALFGBG-71320), the Alzheimer Drug Discovery Foundation (ADDF), USA (#201809-2016862), the AD Strategic Fund and the Alzheimer’s Association (#ADSF-21-831376-C, #ADSF-21-831381-C, and #ADSF-21-831377-C), the Bluefield Project, the Olav Thon Foundation, the Erling-Persson Family Foundation, Stiftelsen för Gamla Tjänarinnor, Hjärnfonden, Sweden (#FO2022-0270), the European Union’s Horizon 2020 research and innovation programme under the Marie Skłodowska-Curie grant agreement No 860197 (MIRIADE), the European Union Joint Programme – Neurodegenerative Disease Research (JPND2021-00694), the National Institute for Health and Care Research University College London Hospitals Biomedical Research Centre, and the UK Dementia Research Institute at UCL (UKDRI-1003). AN reports grants from Parkinson’s UK, Barts Charity, Cure Parkinson’s, National Institute for Health and Care Research, Innovate UK, Virginia Keiley benefaction, Solvemed, the Medical College of Saint Bartholomew’s Hospital Trust, Alchemab, Aligning Science Across Parkinson’s Global Parkinson’s Genetics Program (ASAP-GP2) and the Michael J Fox Foundation. SJT received research grant funding from the Wellcome Trust (223082/Z/21/Z), and the UK Dementia Research Institute that receives its funding from DRI Ltd., funded by the UK MRC, Alzheimer’s Society, and Alzheimer’s Research UK. We are grateful to the United to End MND (U2EM) and UK MND Research Institute consortium for their contributions for the recruitment of the ALS patients in this study.

