## Supplementary Materials for "Validation of remote collection and quantification of blood Neurofilament light in neurological diseases"

### Results

Supplementary Figure 1 – pilot data

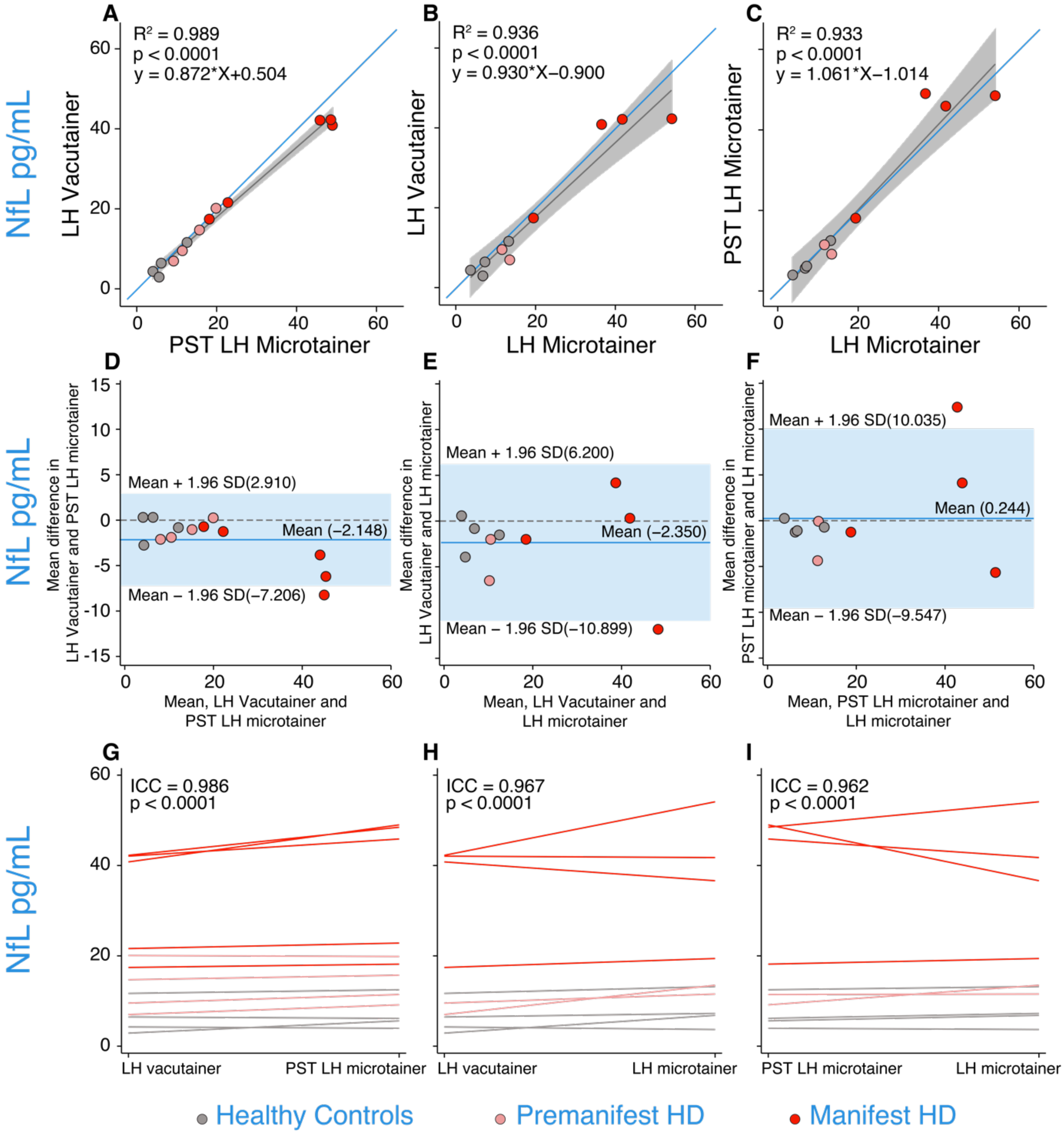


*Supplementary Figure 1: Pilot venous versus capillary tube type comparison data showed our finger-prick method is closer to y=x than previously reported DPS methods (21, 22). Panels A-C are scatter plots of NfL concentrations measured from A) LH vacutainer versus PST LH microtainer, B) LH vacutainer versus LH microtainer, and C) PST LH microtainer versus LH microtainer. Blue lines represent y=x. Grey lines represent the linear regression fit of the data. $R^2$ and P values are generated from regression models comparing the two collection types in each panel. Panels D-F show Bland-*

*Altman plots for agreement between tube-type for NfL measurements with the 95% limits of agreement represented by the limits of the light blue shaded region; dark blue lines represent the mean; dashed line represents y=0; D) LH vacutainer versus PST LH microtainer, E) LH vacutainer versus LH microtainer, F) PST LH microtainer versus LH microtainer. Panels G-I show spaghetti plots to represent the reliability of NfL measurements between G) PST LH microtainer versus LH microtainer, H) LH vacutainer versus LH microtainer, and I) LH microtainer versus PST LH microtainer. NfL, Neurofilament light; LH, Lithium Heparin; PST, Plasma separating tube; HD, Huntington's disease.*

#### Group demographics

*Supplementary Table 1: Group demographics for all combined baseline data followed by each group demographics for each objective.*

| Total | Controls | PD | Pre-HD | HD | ALS | MS | p-value |
|---|---|---|---|---|---|---|---|
| **n** | 40 | 13 | 30 | 29 | 7 | 34 | - |
| **Age (median, IQR)** | 36.4 (29.6 – 58.2) | 66.6 (61.5 – 73.1) | 46 (39.4 – 52.3) | 58.5 (51.5 – 61.5) | 62.8 (49.7 – 76.2) | 57.3 (52 – 60.3) | <0.0005 |
| **Sex (%M)** | 40 | 46 | 13 | 59 | 86 | 38 | 0.0017 |
| **A.0.0** | | | | | | | |
| **n** | 13 | 11 | 6 | 8 | 7 | 13 | - |
| **Age (median, IQR)** | 35.2 (26.9 - 45.8) | 64.8 (53 – 71.8) | 50.1 (36.9 – 59.6) | 60.7 (50.4 – 62.2) | 62.8 (49.7 – 76.2) | 52 (48.3 – 56.8) | 0.0015 |
| **Sex (%M)** | 38 | 45 | 17 | 50 | 86 | 38 | 0.2083 |
| **B.1.a** | | | | | | | |
| **n** | 11 | - | 6 | 5 | - | 5 | - |
| **Age (median, IQR)** | 31 (28.6 – 34.5) | - | 49.2 (46.5 – 53.9) | 51.5 (48 – 58.1) | - | 59.6 (57,5 – 60.3) | 0.0244 |
| **Sex (%M)** | 45 | - | 0 | 80 | - | 60 | 0.0162 |
| **B.1.b** | | | | | | | |
| **n** | 10 | - | 5 | 5 | - | 6 | - |
| **Age (median, IQR)** | 32.1 (30.8 – 61) | - | 42.2 (40.3 – 52.3) | 59.3 (52.5 – 60.3) | - | 59.7 (59.3 – 60.4) | 0.0936 |
| **Sex (%M)** | 40 | - | 20 | 20 | - | 33 | 0.8812 |
| **B.2.a** | | | | | | | |
| **n** | 10 | - | 5 | 5 | - | 5 | - |
| **Age (median, IQR)** | 31.2 (22.9 – 54.9) | - | 39.4 (31.7 – 41.3) | 58.5 (57.8 – 68.9) | - | 59.1 (54.5 – 60.2) | 0.0202 |
| **Sex (%M)** | 50 | - | 0 | 60 | - | 20 | 0.2346 |
| **B.2.b** | | | | | | | |
| **n** | 10 | - | 5 | 6 | - | 5 | - |

| | | | | | | | |
|---|---|---|---|---|---|---|---|
| **Age (median, IQR)** | 32.7 (27.5 – 48.5) | - | 45.4 (44.7 – 54.3) | 57.4 (46.9 – 61.5) | - | 55.2 (55.1 – 66.5) | 0.0136 |
| **Sex (%M)** | 20 | - | 20 | 83 | - | 40 | 0.0638 |

ALS and MS disease subgroups were combined for groupwise comparisons. Data are median (IQR) for age and % male for sex. P-values for age from the Kruskal-Wallis test and for sex from the chi-squared test. HD, Huntington's disease; PD, Parkinson's disease; MS, multiple sclerosis; ALS, amyotrophic lateral sclerosis.

#### Supplementary Figure 2 - A00 paired t-tests

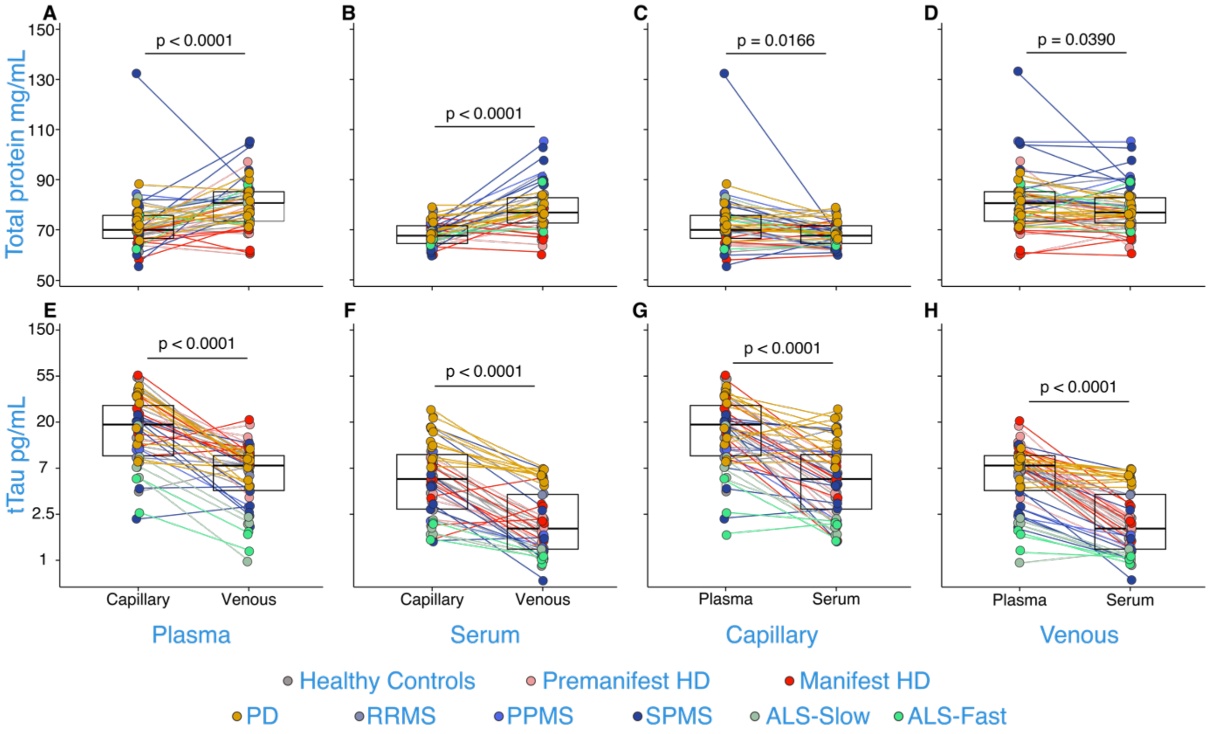


*Supplementary Figure 2: Total protein (A-D) and tTau (E-H) across different collection methods and sample types from experiment A.0.0. p-values were generated from paired t-tests. Boxes represent the interquartile range with bold black horizontal line indicating the median. tTau concentrations are natural log transformed. HD, Huntington's disease; PD, Parkinson's disease; RRMS, relapsing-remitting multiple sclerosis; PPMS, primary progressive multiple sclerosis; SPMS, secondary progressive multiple sclerosis; ALS, amyotrophic lateral sclerosis; tTau, total tau.*

#### Supplementary Figure 3 – A00 Bland-Altman Plots

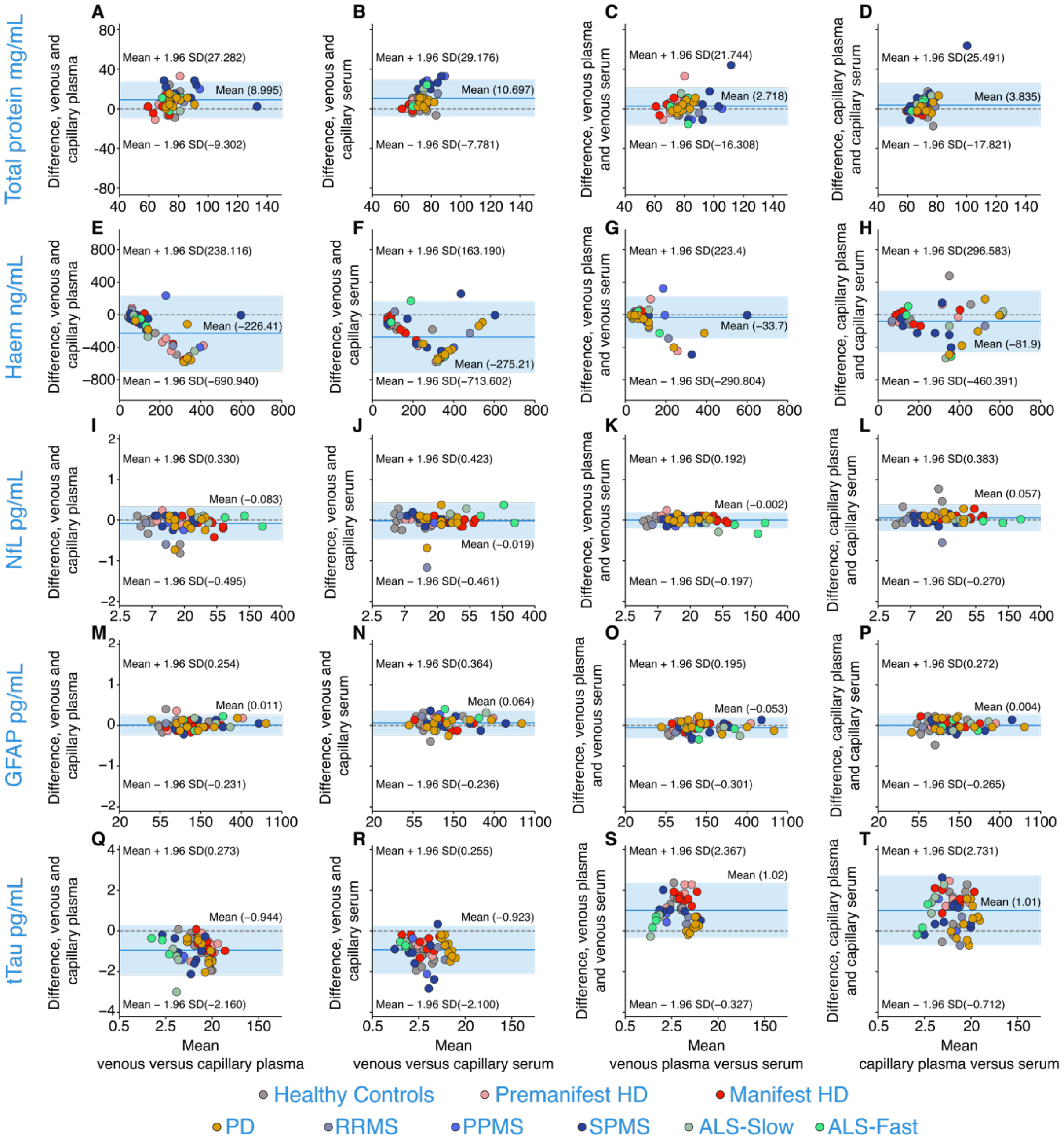


*Supplementary Figure 3: Bland-Altman plots across different collection methods and sample types from experiment A.0.0 for total protein (A-D), haemoglobin (E-H), NfL (I-L), GFAP (M-P), and tTau (Q-T) in healthy controls, pre-HD, manifest-HD, PD, RRMS, PPMS, SPMS, ALS-slow, ALS-fast. 95% limits of agreement are represented by the limits of the light blue shaded region, dark blue lines represent the mean, and the dashed line represents y=0 i.e. no difference NfL, GFAP, and tTau concentrations are natural log transformed. HD, Huntington's disease; PD, Parkinson's disease; RRMS, relapsing-remitting*

*multiple sclerosis; PPMS, primary progressive multiple sclerosis; SPMS, secondary progressive multiple sclerosis; ALS, amyotrophic lateral sclerosis: Haem, haemoglobin; tTau, total tau.*

Supplementary Figure 4 - B1a paired t-tests

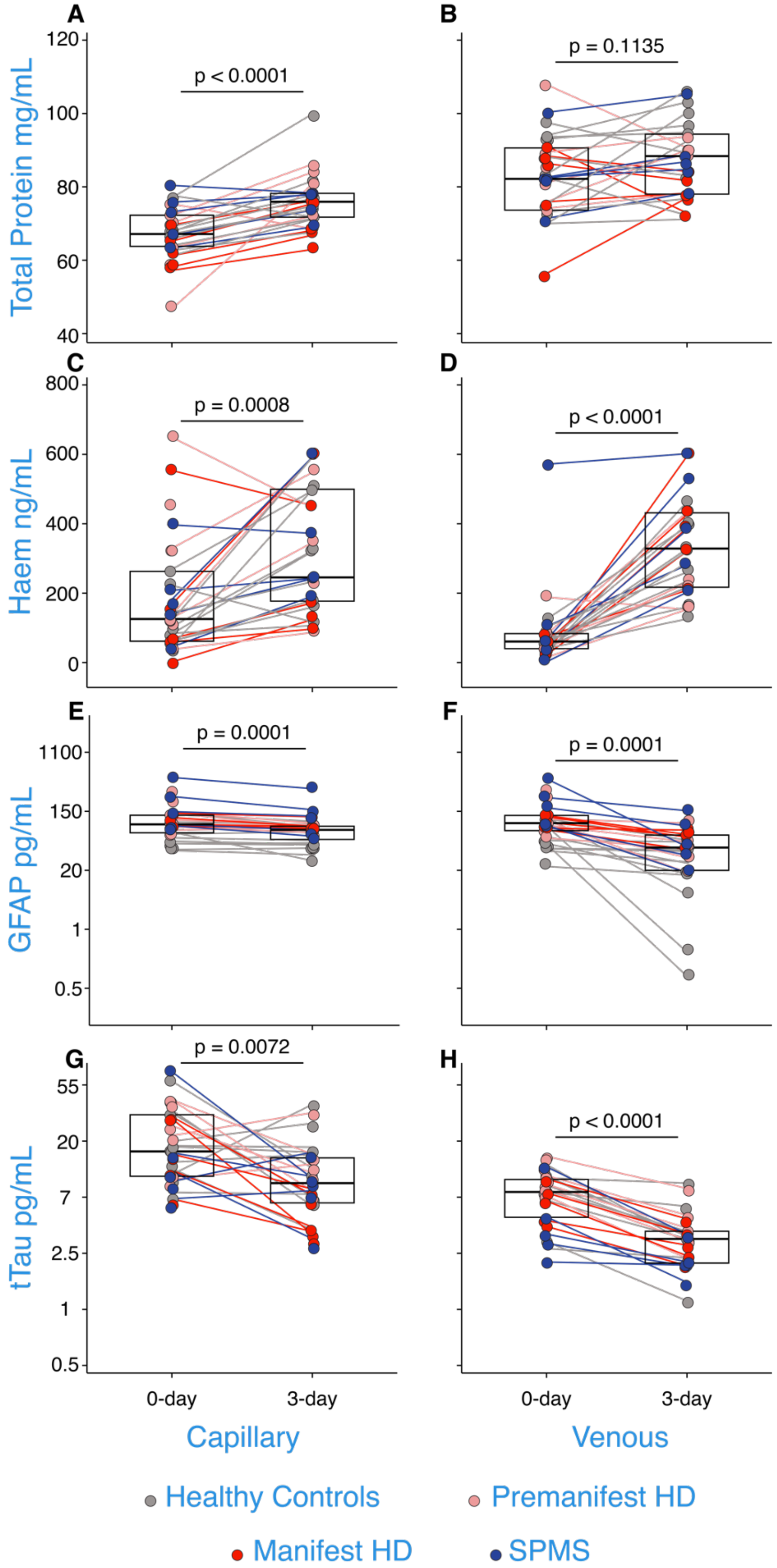


*Supplementary Figure 4: Concentrations of total protein (A-B), haemoglobin (C-D), GFAP (E-F), and tTau (G-H) in capillary and venous plasma when processed on the day of collection and when*

*processed three days later from experiment B.1.a. p-values were generated from paired t-tests. Boxes represent the interquartile range with bold black horizontal line indicating the median. GFAP and tTau concentrations are natural log transformed. HD, Huntington's disease; SPMS, secondary progressive multiple sclerosis; Haem, haemoglobin; tTau, total tau.*

#### Supplementary Figure 5 – B2a paired t-tests

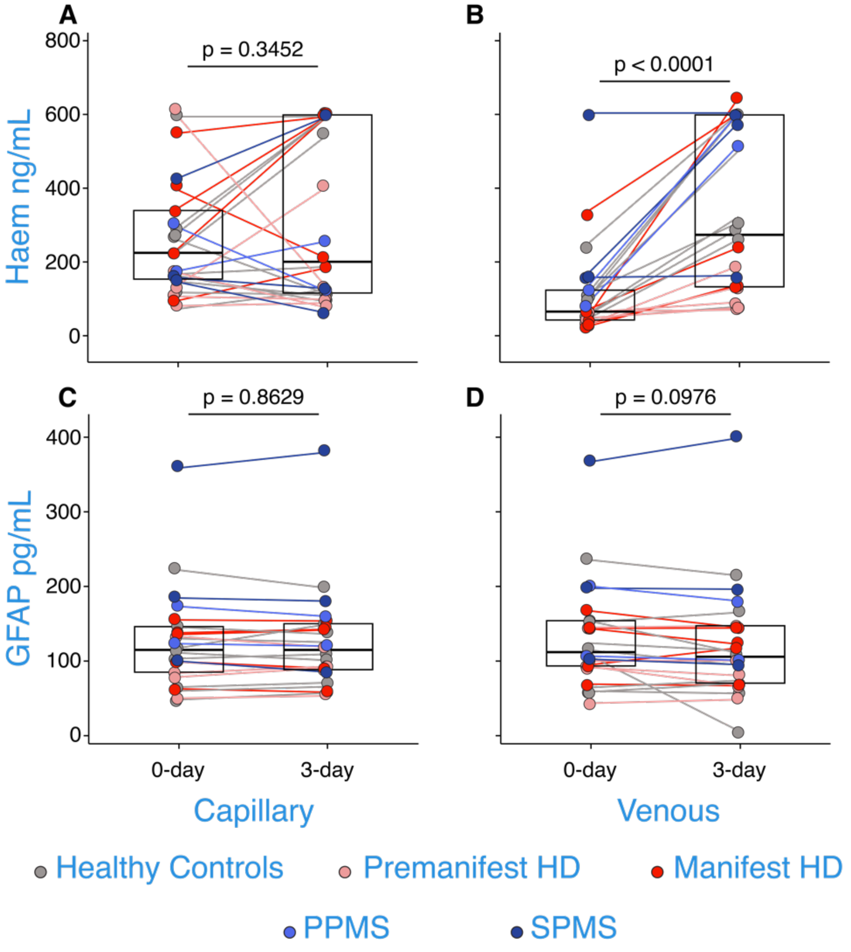


*Supplementary Figure 5: Concentrations of haemoglobin (A-B) and GFAP (C-D) in capillary and venous serum when processed on the day of collection and when processed three days later from experiment B.2.a. p-values were generated from paired t-tests. Boxes represent the interquartile range with bold black horizontal line indicating the median. Haem, haemoglobin; HD, Huntington's disease; PPMS, primary progressive multiple sclerosis; SPMS, secondary progressive multiple sclerosis.*

Supplementary Figure 6 – B1b paired t-tests

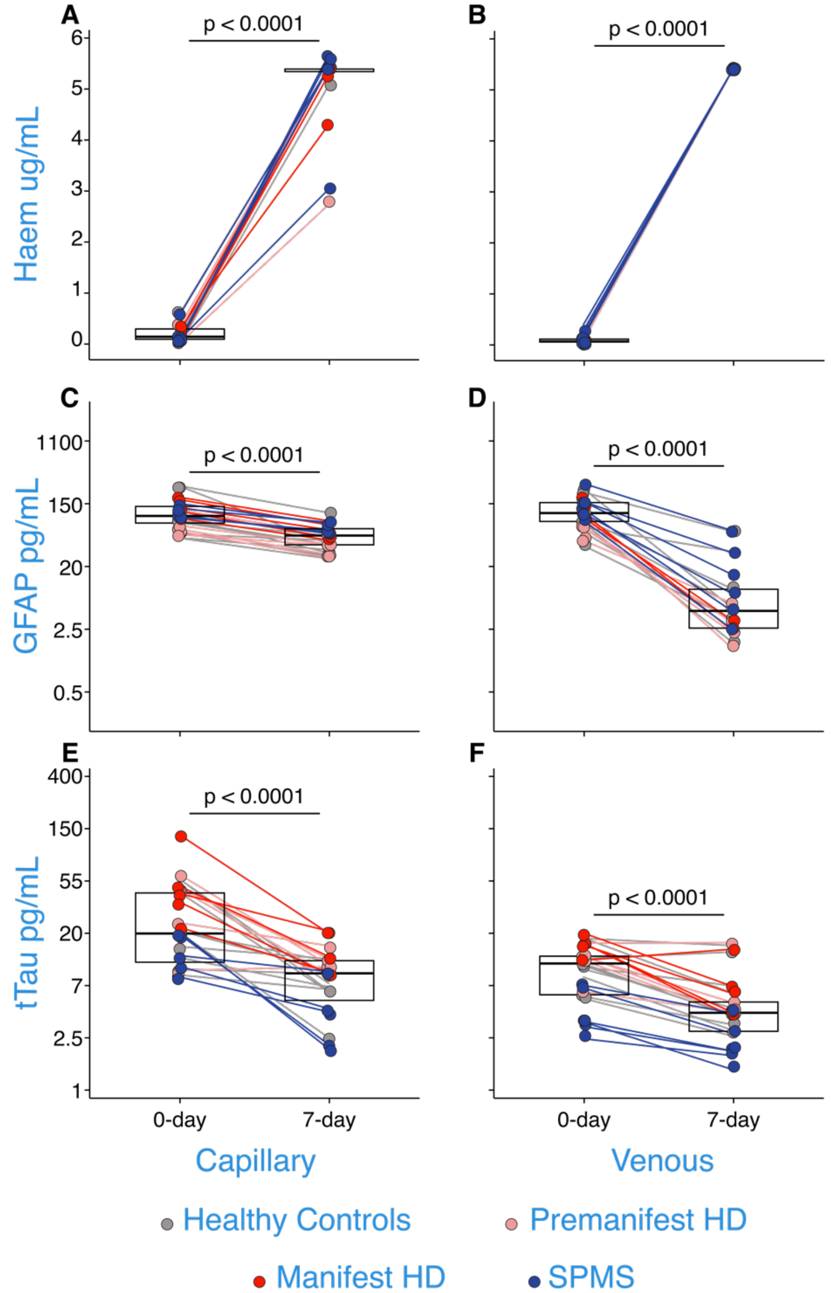


*Supplementary Figure 6: Concentrations of haemoglobin (A-B), GFAP (C-D), and tTau (E-F) from capillary and venous plasma when processed on the day of collection and when processed seven days later from experiment B.1.b. p-values were generated from paired t-tests. Boxes represent the interquartile range with bold black horizontal line indicating the median. GFAP and tTau concentrations are natural log transformed. HD, Huntington's disease; SPMS, secondary progressive multiple sclerosis; Haem, haemoglobin; tTau, total tau.*

#### Supplementary Figure 7 – B2b paired t-tests

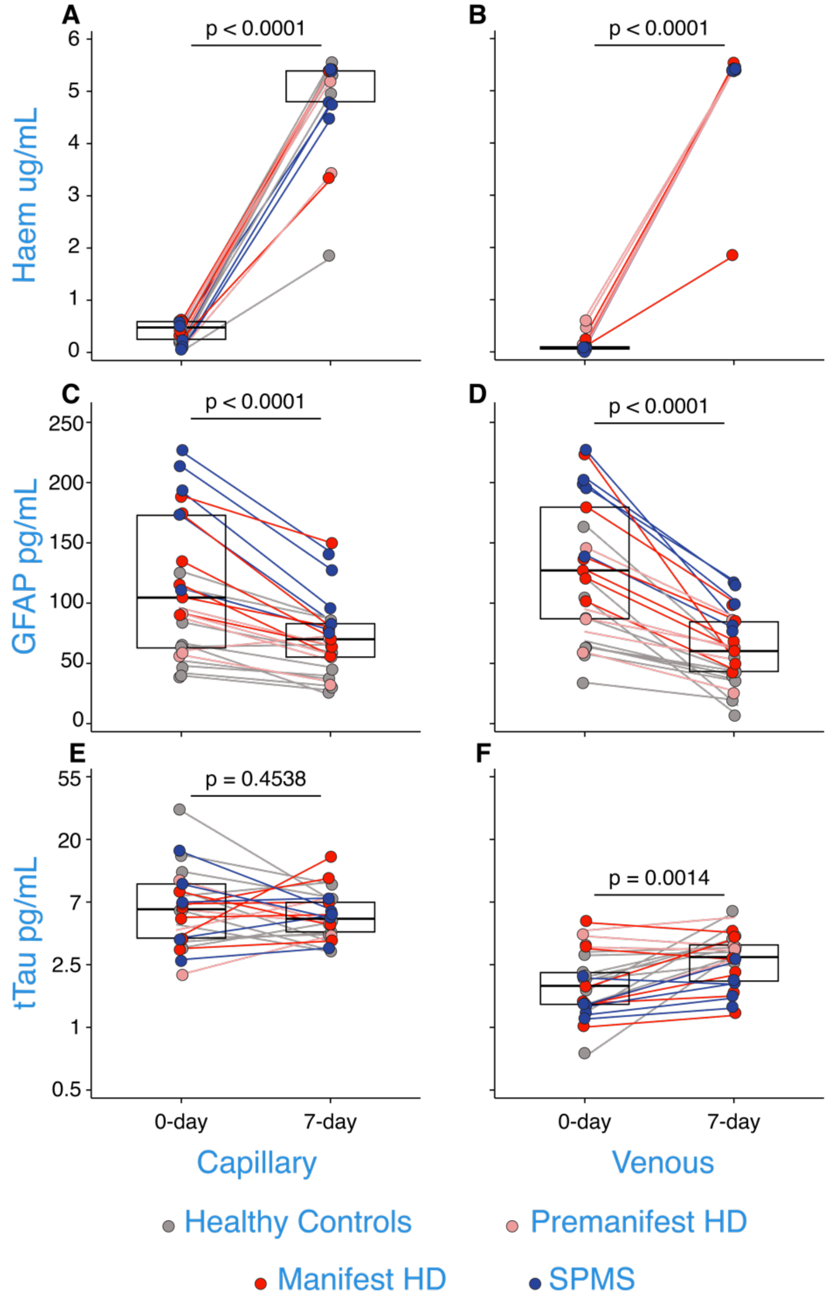


*Supplementary Figure 7: Concentrations of haemoglobin (A-B), GFAP (C-D), and tTau (E-F) from capillary and venous serum when processed on the day of collection and when processed seven days later from experiment B.2.b. p-values were generated from paired t-tests. Boxes represent the interquartile range with bold black horizontal line indicating the median. tTau concentrations are natural log transformed. HD, Huntington's disease; SPMS, secondary progressive multiple sclerosis; Haem, haemoglobin; tTau, total tau.*

*Supplementary Table 2: Mean differences for all baseline NfL data.*

| NfL pg/mL | | PD – controls | | Pre-HD – controls | | Manifest-HD – controls | | ALS – controls | | MS – controls | |
|---|---|---|---|---|---|---|---|---|---|---|---|
| | | (95% CI) | *p* | (95% CI) | *p* | (95% CI) | *p* | (95% CI) | *p* | (95% CI) | *p* |
| Venous | Plasma | 12.07 (0.11,0.83) | 0.010 | **14.65 (0.59, 1.11)** | **< 0.0001** | **38.45 (1.16, 1.73)** | **< 0.0001** | **68.53 (1.2, 2.02)** | **< 0.0001** | 5.28 (-0.23, 0.52) | 0.072 |
| | Serum | 11.66 (-0.01, 0.66) | 0.051 | **9.08 (0.33, 0.89)** | **< 0.0001** | **35.31 (1.01, 1.57)** | **< 0.0001** | **81.03 (1.31, 2.1)** | **< 0.0001** | 5.36 (-0.02, 0.51) | 0.067 |
| Capillary | Plasma | 11.21 (0.06, 0.88) | 0.024 | **12.36 (0.41, 1.01)** | **< 0.0001** | **38.55 (1.05, 1.71)** | **< 0.0001** | **67.98 (1.03, 1.97)** | **< 0.0001** | 2.21 (-0.18, 0.44) | 0.396 |
| | Serum | **11.83 (0.059, 0.76)** | **0.0023** | **7.66 (0.25, 0.88)** | **0.001** | **34.91 (1.02, 1.62)** | **< 0.0001** | **70.22 (1.2, 2.05)** | **< 0.0001** | 5.71 (0.37, 0.6) | 0.027 |

ALS and MS disease subgroups were combined for groupwise comparisons. Data are mean (95% CI), p-value. P-values were generated from multiple linear regressions. The Bonferroni threshold for this experiment was 0.0025 (20 comparisons) and all statistics which reached significance below this are highlighted in bold.CI, confidence interval; NfL, neurofilament light; HD, Huntington's disease; PD, Parkinson's disease; MS, multiple sclerosis; ALS, amyotrophic lateral sclerosis.

*Supplementary Table 3: Mean differences for all baseline GFAP data.*

| GFAP pg/mL | | PD – controls | | Pre-HD – controls | | Manifest-HD – controls | | ALS – controls | | MS – controls | |
|---|---|---|---|---|---|---|---|---|---|---|---|
| | | (95% CI) | *p* | (95% CI) | *p* | (95% CI) | (95% CI) | *p* | (95% CI) | *p* | (95% CI) |
| Venous | Plasma | 103.45 (-0.34, 0.4) | 0.873 | 28.25 (-0.16, 0.37) | 0.441 | 29.55 (-0.21, 0.38) | 0.572 | 83.05 (-0.24, 0.6) | 0.404 | 74.55 (-0.07, 0.48) | 0.148 |
| | Serum | 104.6 (-0.35, 0.12) | 0.916 | 16.4 (-0.23, 0.36) | 0.668 | 31.1 (-0.29, 028) | 0.976 | 98.6 (-0.17, 0.64) | 0.257 | 73.6 (-0.05, 0.49) | 0.111 |
| Capillary | Plasma | 95.7 (-0.35, 0.35) | 0.992 | 19.9 (-0.2, 0.32) | 0.653 | 26.9 (-0.23, 0.34) | 0.695 | 71.5 (-0.3, 0.51) | 0.608 | 57.8 (-0.15, 0.38) | 0.403 |
| | Serum | 103.21 (-0.23, 0.44) | 0.543 | 13.21 (-0.23, 0.37) | 0.625 | 37.91 (-0.19, 0.38) | 0.509 | 77.11 (-0.16, 0.64) | 0.236 | 77.11 (0.04, 0.58) | 0.024 |

ALS and MS disease subgroups were combined for groupwise comparisons. Data are mean (95% CI), p-value. P-values were generated from multiple linear regressions. The Bonferroni threshold for this experiment was 0.0025 (20 comparisons) and all statistics which reached significance below this are highlighted in bold. CI, confidence interval; NfL, neurofilament light; HD, Huntington's disease; PD, Parkinson's disease; MS, multiple sclerosis; ALS, amyotrophic lateral sclerosis.

Supplementary Figure 8 – GFAP group comparisons for all baseline data

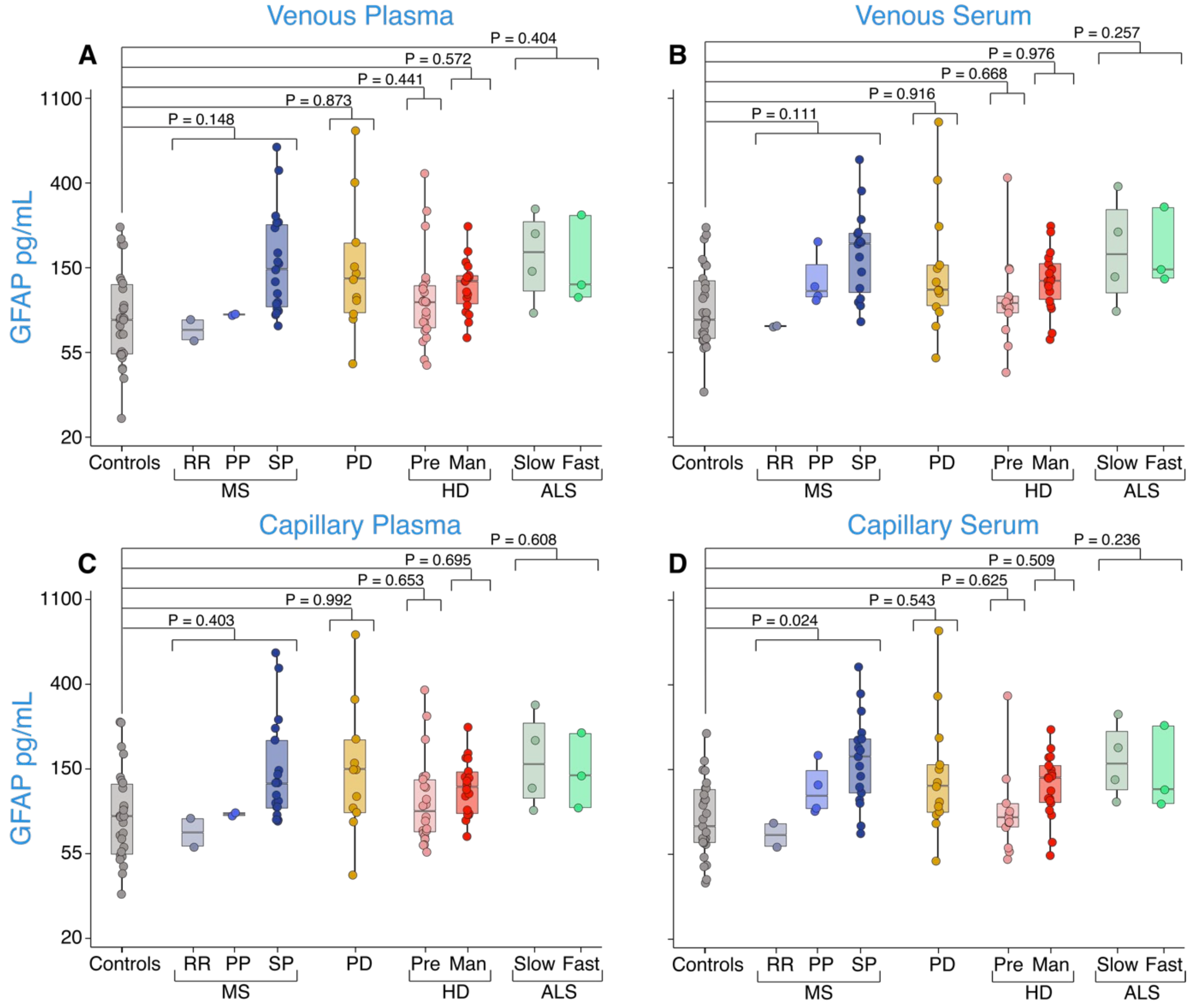


*Supplementary Figure 8: Disease group comparison of GFAP concentrations from venous plasma (A), venous serum (B), capillary plasma (C), and capillary serum (D) from all baseline data for healthy controls, PPMS, RRMS, SPMS, PD, pre-HD, manifest-HD, ALS-slow and ALS-fast patients. P values were generated from multiple linear regressions. The Bonferroni threshold for this experiment was 0.0025 (20 comparisons) and all statistics which reached significance below this are highlighted in bold. GFAP concentrations are natural log transformed. PD, Parkinson's disease; RR, relapsing-remitting; PP, primary progressive; SP, secondary progressive; MS, multiple sclerosis; HD, Huntington's disease; Pre, premanifest; Man, manifest; ALS, amyotrophic lateral sclerosis.*

*Supplementary Table 4: Summary measures for NfL pg/mL from each objective.*

| NfL pg/mL | | Controls | PD | Pre-HD | Manifest-HD | ALS | MS |
|---|---|---|---|---|---|---|---|
| **A.0.0** | | | | | | | |
| Venous | Plasma | 10.07 (5.76) | 22.29 (10.41) | 20.19 (9.39) | 42.8 (13.5) | 78.75 (64.19) | 13.8 (6.86) |
| | Serum | 9.79 (5.39) | 21.82 (9.85) | 18.9 (9.91) | 42.67 (15.09) | 91.19 (75.73) | 14.02 (6.57) |
| Capillary | Plasma | 12.69 (8.38) | 24.09 (10.59) | 20.29 (9.88) | 48.84 (18.32) | 80.86 (74.72) | 15.13 (6.92) |
| | Serum | 10.57 (5.93) | 22.32 (9.56) | 17.43 (9.64) | 44.72 (16.14) | 80.71 (71.89) | 15.79 (7.53) |
| **B.1.a** | | | | | | | |
| Venous | Plasma day 0 | 7.36 (4.03) | - | 24.09 (7.76) | 52.48 (13.63) | - | 17.07 (7.35) |
| | Plasma day 3 | 7.13 (3.64) | - | 24.53 (8.42) | 53.28 (14.75) | - | 15.89 (7.39) |
| Capillary | Plasma day 0 | 10.38 (10.41) | - | 24.9 (8.73) | 53.28 (13.28) | - | 17.77 (7.04) |
| | Plasma day 3 | 7.76 (4.14) | - | 32.73 (9.49) | 54.33 (15.81) | - | 18.66 (8.42) |
| **B.1.b** | | | | | | | |
| Venous | Plasma day 0 | 9.93 (5.58) | - | 31.91 (21.55) | 54.25 (27.58) | - | 17.59 (8.67) |
| | Plasma day 7 | 10.51 (6.38) | - | 27.82 (20.91) | 45.64 (22.42) | - | 18.25 (11.32) |
| Capillary | Plasma day 0 | 10.97 (5.48) | - | 31.79 (24.51) | 53.71 (25.55) | - | 12.29 (3.52) |
| | Plasma day 7 | 10.08 (5.23) | - | 31.33 (24.77) | 48.93 (25.33) | - | 12.01 (3.12) |
| **B.2.a** | | | | | | | |
| Venous | Serum day 0 | 10.38 (5.21) | - | 21.18 (10.04) | 45.38 (13.75) | - | 17.5 (4.76) |
| | Serum day 3 | 10.44 (5.7) | - | 22.37 (11.59) | 47.35 (13.93) | - | 17.71 (4.31) |
| Capillary | Serum day 0 | 10.31 (4.79) | - | 19.52 (8.73) | 43.2 (11.85) | - | 16.48 (4.72) |
| | Serum day 3 | 10.85 (4.88) | - | 21.82 (9.84) | 43.76 (11.61) | - | 17.11 (5.04) |
| **B.2.b** | | | | | | | |
| Venous | Serum day 0 | 7.92 (3.74) | - | 19.15 (9.26) | 49.29 (27.22) | - | 17.15 (5.76) |
| | Serum day 7 | 7.42 (3.37) | - | 19.23 (8.83) | 42.4 (23.76) | - | 15.91 (4.62) |
| Capillary | Serum day 0 | 7.64 (5.43) | - | 19.97 (9.2) | 48.15 (26.06) | - | 16.97 (4.63) |
| | Serum day 7 | 8.49 (7.64) | - | 18.48 (8.99) | 43.82 (24.17) | - | 15.7 (3.62) |

Data are mean (SD). HD, Huntington's disease; PD, Parkinson's disease; MS, multiple sclerosis; ALS, amyotrophic lateral sclerosis.

*Supplementary Table 2: Summary measures for GFAP pg/mL from each objective.*

| GFAP pg/mL | | Controls | PD | Pre-HD | Manifest-HD | ALS | MS |
|---|---|---|---|---|---|---|---|
| **A.0.0** | | | | | | | |
| Venous | Plasma | 97 (49.54) | 199.3 (205.9) | 153.5 (150.5) | 129.6 (55.51) | 178.9 (85.12) | 160.6 (160.1) |
| | Serum | 103.2 (53.79) | 208.3 (230.8) | 152.8 (139.7) | 131.1 (53.77) | 202.3 (108.4) | 163.1 (132.9) |
| Capillary | Plasma | 95.58 (47.14) | 194.5 (193.5) | 137.7 (121.2) | 135.1 (55.36) | 170.3 (82.89) | 154 (143.8) |
| | Serum | 90.66 (37.31) | 194.4 (207.5) | 140.6 (122.9) | 131 (57.6) | 168.3 (74.98) | 154 (124.6) |
| **B.1.a** | | | | | | | |
| Venous | Plasma day 0 | 66.42 (34.42) | - | 134.2 (78.21) | 109.8 (23.54) | - | 215.8 (152.1) |
| | Plasma day 3 | 27.72 (20.4) | - | 51.87 (33.1) | 66.27 (14.62) | - | 71.25 (56.61) |
| Capillary | Plasma day 0 | 66.55 (34.96) | - | 130.3 (71.27) | 99.73 (21.39) | - | 206.4 (167.1) |
| | Plasma day 3 | 54.21 (24.9) | - | 79.13 (9.92) | 85.82 (16.7) | - | 142.5 (105.5) |
| **B.1.b** | | | | | | | |
| Venous | Plasma day 0 | 95.8 (70.56) | - | 70.41 (25.18) | 134.2 (32.76) | - | 152.2 (65.98) |
| | Plasma day 7 | 12.29 (19.23) | - | 2.77 (2.37) | 1.27 (1.77) | - | 20.56 (22.33) |
| Capillary | Plasma day 0 | 107.7 (80.68) | - | 75.12 (25.88) | 136.7 (35.46) | - | 113.5 (21.68) |
| | Plasma day 7 | 44.27 (23.69) | - | 37.94 (11.32) | 63.15 (15.8) | - | 66.59 (11.19) |
| **B.2.a** | | | | | | | |
| Venous | Serum day 0 | 114.2 (57.03) | - | 94.55 (36.34) | 124.2 (40.87) | - | 195.9 (108) |
| | Serum day 3 | 93.15 (62.88) | - | 88.82 (38.29) | 119.6 (31.98) | - | 195.5 (124.5) |
| Capillary | Serum day 0 | 109.9 (51.37) | - | 89.57 (30.69) | 119.1 (37.36) | - | 189.7 (102.6) |
| | Serum day 3 | 106.3 (48.77) | - | 89.23 (23.57) | 118.3 (41.6) | - | 186.8 (116.2) |
| **B.2.b** | | | | | | | |
| Venous | Serum day 0 | 82.71 (37.71) | - | 92.35 (32.74) | 148.5 (45.38) | - | 192.9 (32.19) |
| | Serum day 7 | 33.11 (14.88) | - | 57.8 (21.83) | 67.03 (21.27) | - | 97.64 (17.92) |
| Capillary | Serum day 0 | 69.61 (29.57) | - | 77.71 (18.7) | 135 (39.46) | - | 184.1 (45.18) |
| | Serum day 7 | 48.96 (23.47) | - | 54.94 (14.54) | 82.83 (33.87) | - | 104.2 (28.06) |

Data are mean (SD). HD, Huntington’s disease; PD, Parkinson’s disease; MS, multiple sclerosis; ALS, amyotrophic lateral sclerosis.

*Supplementary Table 6: Summary measures for tTau pg/mL from each objective.*

| tTau pg/mL | | Controls | PD | Pre-HD | Manifest-HD | ALS | MS |
|---|---|---|---|---|---|---|---|
| **A.0.0** | | | | | | | |
| Venous | Plasma | 8.26 (1.62) | 7.94 (2.01) | 12.47 (4.87) | 11.48 (4.2) | 1.84 (0.56) | 5.05 (3.13) |
| | Serum | 2.13 (1.38) | 6.3 (0.72) | 1.94 (0.51) | 2.71 (1.51) | 1.08 (0.1) | 2.34 (1.94) |
| Capillary | Plasma | 25.33 (16.24) | 25.97 (13.67) | 25.41 (9.33) | 25.01 (15.19) | 7.5 (5.74) | 14.43 (6.98) |
| | Serum | 6.49 (2.41) | 15.3 (6.34) | 4.35 (2.91) | 5.08 (2.48) | 2 (0.31) | 8.07 (6) |
| **B.1.a** | | | | | | | |
| Venous | Plasma day 0 | 7.64 (3.39) | - | 10.39 (2.61) | 6.7 (2.28) | - | 5.33 (4.03) |
| | Plasma day 3 | 3.89 (2.09) | - | 4.6 (2.25) | 3.22 (1.06) | - | 2.34 (0.76) |
| Capillary | Plasma day 0 | 20.11 (15.96) | - | 25.96 (13.86) | 18.36 (10.93) | - | 22.24 (27.22) |
| | Plasma day 3 | 13.14 (10.53) | - | 15.02 (9.59) | 5.01 (1.93) | - | 8.78 (4.4) |
| **B.1.b** | | | | | | | |
| Venous | Plasma day 0 | 11.23 (4.17) | - | 12.22 (3.53) | 16.17 (2.82) | - | 4.8 (2.04) |
| | Plasma day 7 | 6.69 (4.55) | - | 7.12 (5.31) | 7.43 (4.31) | - | 2.6 (1.04) |
| Capillary | Plasma day 0 | 25.74 (17.87) | - | 37.12 (20.08) | 55.31 (42.4) | - | 13.94 (5.01) |
| | Plasma day 7 | 7.83 (2.76) | - | 11.41 (2.55) | 14.14 (5.57) | - | 4.52 (2.95) |
| **B.2.a** | | | | | | | |
| Venous | Serum day 0 | 2.2 (0.99) | - | 2.53 (0.81) | 2.37 (1.49) | - | 2 (1.26) |
| | Serum day 3 | 3.4 (1.27) | - | 3.36 (1.01) | 2.86 (1.52) | - | 2.34 (1.21) |
| Capillary | Serum day 0 | 8.86 (4.87) | - | 9.27 (4.19) | 7.83 (4.25) | - | 7.59 (6.21) |
| | Serum day 3 | 11.5 (8.92) | - | 15.63 (10.58) | 4.16 (1.82) | - | 7.56 (4.24) |
| **B.2.b** | | | | | | | |
| Venous | Serum day 0 | 2.1 (0.85) | - | 3.12 (1.7) | 2.52 (1.71) | - | 1.52 (0.43) |
| | Serum day 7 | 4.02 (1.1) | - | 5.03 (2.41) | 2.89 (1.37) | - | 2.02 (0.58) |
| Capillary | Serum day 0 | 10.06 (8.79) | - | 7.73 (4.85) | 6.02 (1.97) | - | 8.21 (5.51) |
| | Serum day 7 | 5.99 (2.57) | - | 8.14 (5.59) | 8.07 (4.22) | - | 5.9 (1.54) |

Data are mean (SD). HD, Huntington's disease; PD, Parkinson's disease; MS, multiple sclerosis; ALS, amyotrophic lateral sclerosis; tTau, total tau.

*Supplementary Table 7: Summary measures for total protein mg/mL from each objective.*

| Total protein mg/mL | | Controls | PD | Pre-HD | Manifest-HD | ALS | MS |
|---|---|---|---|---|---|---|---|
| **A.0.0** | | | | | | | |
| Venous | Plasma | 76.79 (6.267) | 80.55 (6.88) | 78.64 (12.89) | 72.63 (8.91) | 81.27 (5.9) | 93.67 (15.96) |
| | Serum | 76.2 (5.41) | 77.82 (3.35) | 72.39 (5.93) | 70.16 (6.35) | 78.96 (7.27) | 90.01 (8.812) |
| Capillary | Plasma | 69.51 (5.28) | 74.02 (6.19) | 67.5 (4.12) | 67.67 (5.54) | 73.71 (6.51) | 76.98 (18.77) |
| | Serum | 69.33 (7.56) | 71.63 (3.97) | 65.75 (3.47) | 68.11 (5.68) | 66.53 (1.99) | 67.15 (4.96) |
| **B.1.a** | | | | | | | |
| Venous | Plasma day 0 | 83.11 (10.63) | - | 74.33 (19.56) | 79.12 (14.38) | - | 83.42 (10.51) |
| | Plasma day 3 | 89.31 (12.51) | - | 87.91 (5.99) | 78.29 (4.46) | - | 88.4 (10.06) |
| Capillary | Plasma day 0 | 68.24 (5.04) | - | 62.74 (12.22) | 62.83 (4.96) | - | 72.11 (6.68) |
| | Plasma day 3 | 76.75 (8.77) | - | 78.34 (7.42) | 70.23 (5.53) | - | 75.52 (3.89) |
| **B.1.b** | | | | | | | |
| Venous | Plasma day 0 | 78.41 (19.14) | - | 86.65 (6.67) | 81.73 (9.5) | - | 81.38 (5.26) |
| | Plasma day 7 | 80.09 (8.08) | - | 82.59 (17.24) | 77.58 (11.04) | - | 85.07 (7.2) |
| Capillary | Plasma day 0 | 67.84 (4.1) | - | 68.37 (4.06) | 64.57 (8.99) | - | 71.78 (5.75) |
| | Plasma day 7 | 65.14 (5.63) | - | 67.79 (6.98) | 64.94 (7.14) | - | 70.14 (3.83) |
| **B.2.a** | | | | | | | |
| Venous | Serum day 0 | 93.54 (15.98) | - | 78.7 (10.38) | 89.95 (9.15) | - | 83.4(8.62) |
| | Serum day 3 | 92.41 (20.07) | - | 106.2 (16.9) | 88.66 (8.9) | - | 88.46 (12.99) |
| Capillary | Serum day 0 | 63.25 (8.99) | - | 62.16 (6.23) | 72.61 (1.56) | - | 69.91 (5.42) |
| | Serum day 3 | 66.44 (5.76) | - | 65.44 (7.08) | 62.57 (6.05) | - | 70.54 (2.68) |
| **B.2.b** | | | | | | | |
| Venous | Serum day 0 | 78.44 (10.15) | - | 89.25 (14.24) | 77.55 (9.26) | - | 91.07 (10.73) |
| | Serum day 7 | 72.08 (10.4) | - | 82.52 (12.9) | 79.09 (10.42) | - | 92.96 (13.95) |
| Capillary | Serum day 0 | 63.14 (8.6) | - | 62.78 (4.35) | 61.13 (7.02) | - | 69.48 (4.08) |
| | Serum day 7 | 58.38 (12.38) | - | 56.22 (3.61) | 57.42 (3.92) | - | 68.2 (4.44) |

Data are mean (SD). HD, Huntington's disease; PD, Parkinson's disease; MS, multiple sclerosis; ALS, amyotrophic lateral sclerosis.

*Supplementary Table 8: Summary measures for haemoglobin ng/mL from each objective.*

| Haemoglobin ng/mL | | Controls | PD | Pre-HD | Manifest-HD | ALS | MS |
|---|---|---|---|---|---|---|---|
| **A.0.0** | | | | | | | |
| Venous | Plasma | 56.35 (16.88) | 64.88 (71.54) | 91.9 (70.22) | 72.26 (31.82) | 66.43 (44.16) | 143.5 (169.8) |
| | Serum | 61.06 (44.82) | 157.3 (163.7) | 112.9 (182.5) | 55.24 (18.38) | 102.5 (81.91) | 193.5 (206) |
| Capillary | Plasma | 296.4 (243.6) | 505.1 (180.1) | 318.3 (231.2) | 211.7 (195.6) | 284.5 (233.4) | 235 (204.8) |
| | Serum | 304.9 (222.4) | 583.7 (49.78) | 306.8 (200.1) | 198.7 (116.9) | 439.6 (228.1) | 380.2 (200.5) |
| **B.1.a** | | | | | | | |
| Venous | Plasma day 0 | 64.94 (29.2) | - | 73.73 (51.03) | 45.78 (21.24) | - | 158.3 (232.8) |
| | Plasma day 3 | 296.4 (103) | - | 243.2 (114.5) | 394.1 (141.5) | - | 403.2 (162.2) |
| Capillary | Plasma day 0 | 110.9 (75.53) | - | 244.9 (208.6) | 169.1 (225.8) | - | 193 (131.6) |
| | Plasma day 3 | 283.2 (184.8) | - | 365 (215.1) | 291.5 (223.1) | - | 332.2 (163.8 |
| **B.1.b** | | | | | | | |
| Venous | Plasma day 0 | 121.6 (173.6) | - | 62.88 (31.99) | 80.16 (30.33) | - | 111 (93.68) |
| | Plasma day 7 | 5400 (0) | - | 5400 (0) | 5400 (0) | - | 5400 (0) |
| Capillary | Plasma day 0 | 222.3 (215.4) | - | 272.2 (214.8) | 175.7 (122.2) | - | 206.2 (222.3) |
| | Plasma day 7 | 4120 (1684) | - | 4873 (1178) | 5153 (479.9) | - | 5017 (1098) |
| **B.2.a** | | | | | | | |
| Venous | Serum day 0 | 89.25 (60.34) | - | 46.32 (16.4) | 98.34 (131.1) | - | 224.3 (212.4) |
| | Serum day 3 | 399.5 (177.7) | - | 110 (49.67) | 349.1 (251.7) | - | 489.7 (187) |
| Capillary | Serum day 0 | 233.7 (146.6) | - | 223.9 (223.2) | 324.5 (174.4) | - | 244.1 (118.8) |
| | Serum day 3 | 310.5 (240) | - | 162.8 (138.4) | 440.2 (219) | - | 235.8 (215.8) |
| **B.2.b** | | | | | | | |
| Venous | Serum day 0 | 53.43 (17.45) | - | 260 (261.1) | 97.95 (81.28) | - | 66 (36.59) |
| | Serum day 7 | 5300 (334.1) | - | 5400 (0) | 4827 (1455) | - | 5400 (0) |
| Capillary | Serum day 0 | 328.6 (216) | - | 355.4 (233.8) | 489.9 (145) | - | 304.7 (233.4) |
| | Serum day 7 | 4420 (1575) | - | 4690 (910.2) | 5058 (838.8) | - | 4963 (414.2) |

Data are mean (SD). HD, Huntington's disease; PD, Parkinson's disease; MS, multiple sclerosis; ALS, amyotrophic lateral sclerosis.

Supplementary Figure 9 – B1a and B2a 3-day delay Bland Altman plots

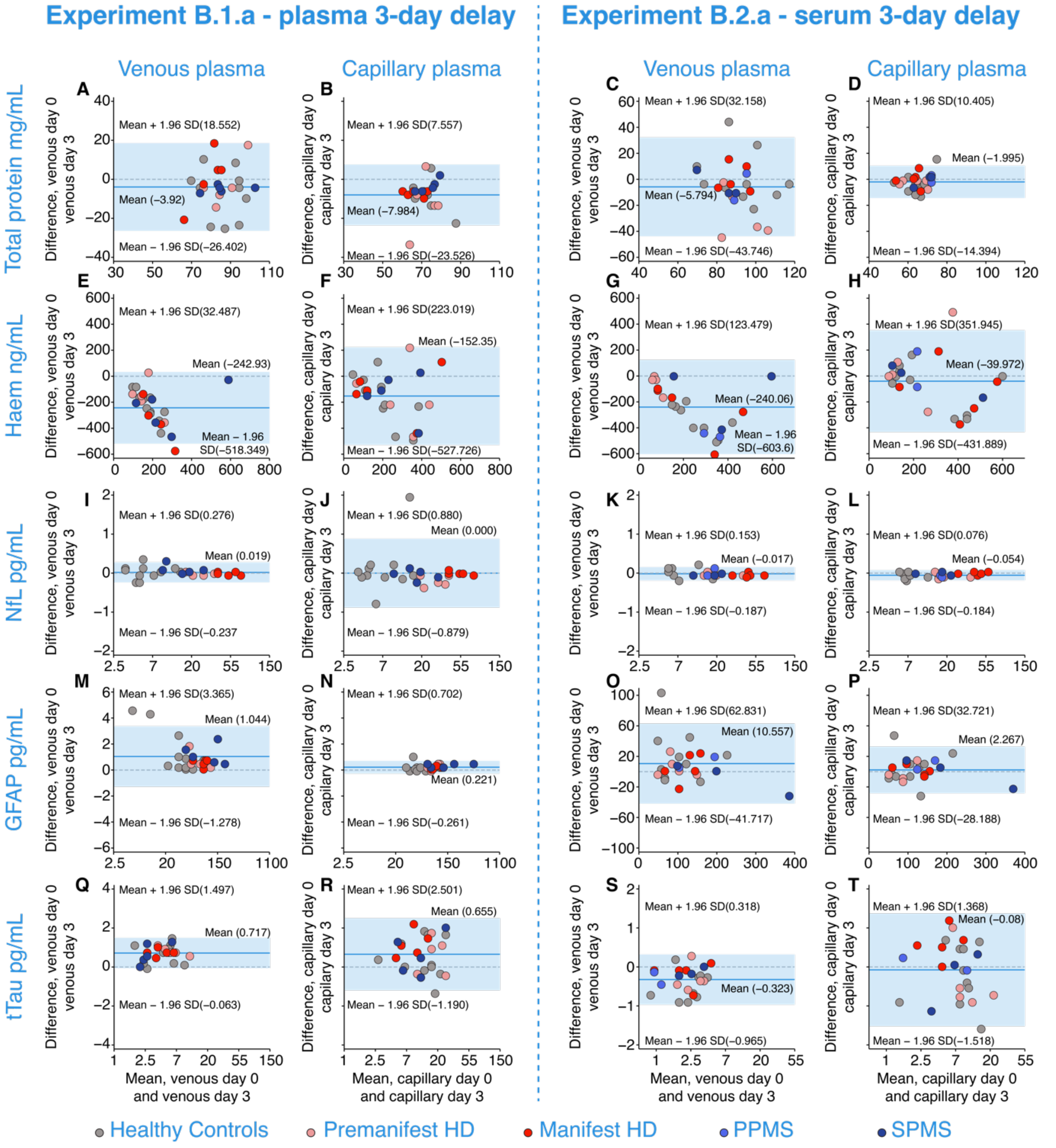


*Supplementary Figure 9: Bland-Altman plots of mean differences of analyte concentrations between zero-day and three-day delay in processing from experiment B.1.a and B.2.a for total protein (A-B), haemoglobin (E-F), NfL (I-J), GFAP (M-N), and tTau (Q-R) and B.2.a for total protein (C-D), haemoglobin (G-H), NfL (K-L), GFAP (O-P), and tTau (S-T) in healthy controls, pre-HD, manifest-HD, PPMS, SPMS. 95% limits of agreement are represented by the limits of the light blue shaded region, dark blue lines represent the mean, and the dashed line represents y=0. NfL, GFAP (only B.1.a (M-N)), and tTau concentrations are natural log transformed. HD, Huntington's disease; PPMS, primary*

*progressive multiple sclerosis; SPMS, secondary progressive multiple sclerosis; Haem, haemoglobin; tTau, total tau.*

#### Supplementary Figure 10 – B1b and B2b 7-day delay Bland Altman plots

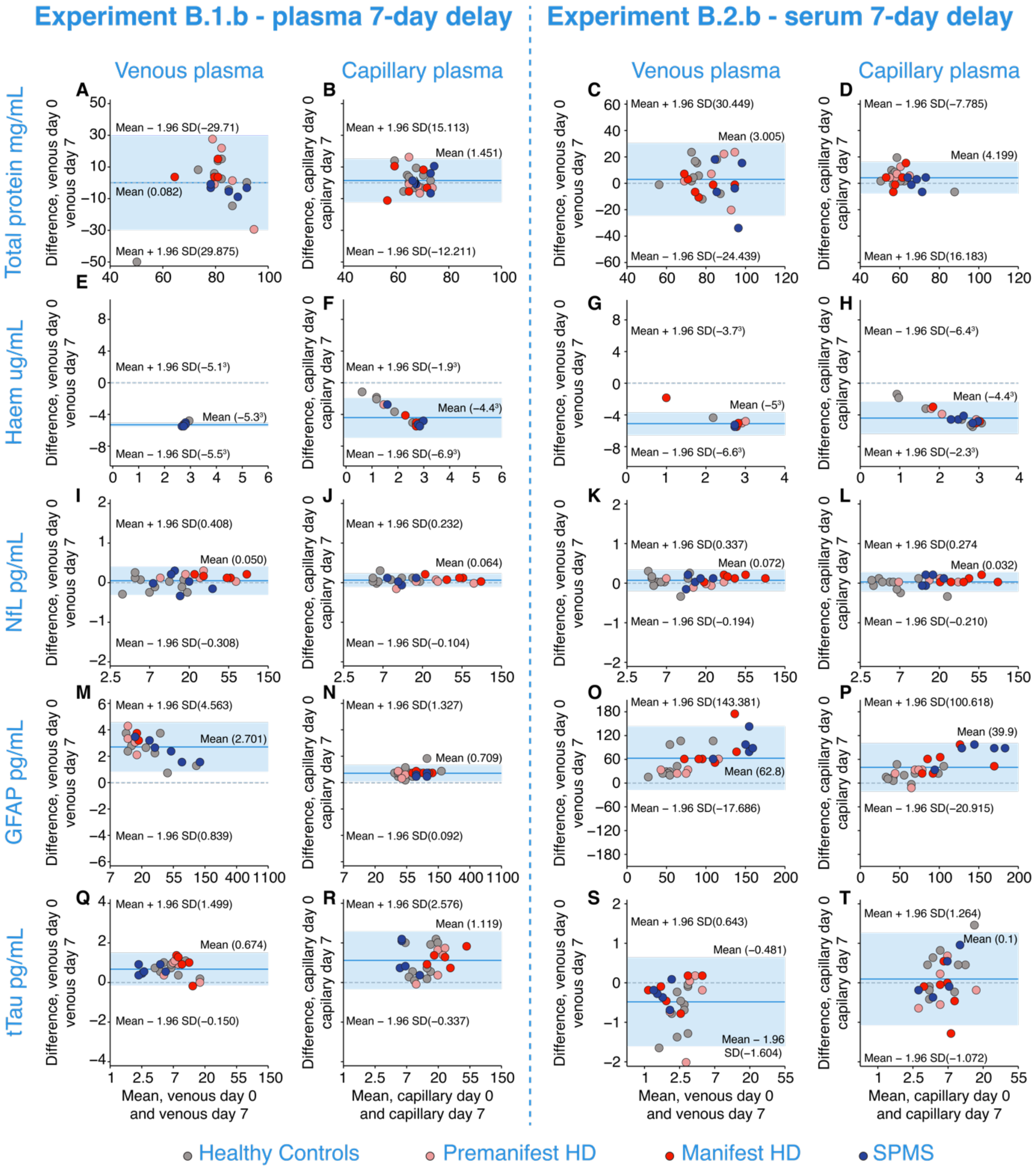


*Supplementary Figure 10: Bland-Altman plots of mean differences of analyte concentrations between zero-day and seven-day delay in processing from experiment B.1.b and B.2.b for total protein (A-B), haemoglobin (E-F), NfL (I-J), GFAP (M-N), and tTau (Q-R) and B.2.b for total protein (C-D), haemoglobin (G-H), NfL (K-L), GFAP (O-P), and tTau (S-T) in healthy controls, pre-HD, manifest-HD, PPMS, SPMS. 95% limits of agreement are represented by the limits of the light blue shaded region,*

*dark blue lines represent the mean, and the dashed line represents y=0. NfL, GFAP (only B.1.b (M-N)), and tTau concentrations are natural log transformed. HD, Huntington's disease; PPMS, primary progressive multiple sclerosis; SPMS, secondary progressive multiple sclerosis; Haem, haemoglobin; tTau, total tau.*

#### Methods

Inter- and Intra- assay Coefficients of Variance (CV) for each analyte measured:

*Supplementary Table 9: Assessment of technical variation for each analyte with Inter-assay % CV and average intra-assay % CV for each analyte measured.*

| Analyte | Inter-assay %CV | Intra-assay %CV |
|---|---|---|
| **NfL** | 16.22 | 4.33 |
| **GFAP** | 19.82 | 7.62 |
| **Tau** | 13.71 | 4.25 |
| **Haemoglobin** | 80.3 | 5.91 |
| **Total protein** | 23.3 | 2.8 |
